# Benchmarking CRISPR-BP34 for point-of-care melioidosis detection in LMIC: a molecular diagnostics study

**DOI:** 10.1101/2023.05.06.23289616

**Authors:** Sukripong Pakdeerat, Phumrapee Boonklang, Kesorn Angchagun, Chalita Chomkatekaew, Yaowaret Dokket, Areeya Faosap, Gumphol Wongsuwan, Vanaporn Wuthiekanun, Panatda Aramrueung, Phadungkiat Khamnoi, Hathairat Thananchai, Suwattiya Siriboon, Parinya Chamnan, Sharon J Peacock, Nicholas PJ Day, Nicholas R Thomson, Chayasith Uttamapinant, Somsakul Pop Wongpalee, Claire Chewapreecha

## Abstract

**Background:** Melioidosis is a grossly neglected but often-fatal tropical disease. The disease is named “a great mimicker” after its broad clinical manifestations, which makes disease diagnosis challenging and time-consuming. To improve diagnosis, we developed and evaluated the performance of the CRISPR-Cas12a system called “CRISPR-BP34” to detect *Burkholderia pseudomallei* DNA across clinical specimens from patients suspected to have melioidosis.

**Methods:** We documented time taken for diagnosis, antibiotics prescribed during the waiting period, and infection outcomes in 875 melioidosis patients treated in a hospital in northeast Thailand between October 2019 and December 2022. In the last six months, we performed CRISPR-BP34 detection on clinical specimens (blood, urine, respiratory secretion, pus and other body fluids) collected from 330 patients with suspected melioidosis and compared its performance to the current gold-standard culture-based method. Discordant results were validated by three independent qPCR tests.

**Findings:** A window of 3-4 days was required for gold-standard culture diagnosis, which resulted in delayed treatment. 199 [22·7%] of 875 patients died prior to diagnosis results while 114 [26·3%] of 433 follow-up cases had been diagnosed, treated, but died within 28 days of admission. A shorter sample-to-diagnosis time of less than 4 hours offered by CRISPR-BP34 technology could lead to faster administration of correct treatment. We demonstrated an improved sensitivity of CRISPR-BP34 (106 [93·0%] of 114 positive cases, 95% CI 86·6 - 96·9) compared to the culture approach (76 [66·7%] of 114 positive cases, 95% CI 57·2 - 75·2); while maintaining similar specificity (209 [96·8%] of 216 negative cases, 95% CI 93·4-98·7) to the culture (216 [100 %] of 216 negative cases, 95% CI 98·3-100·0).

**Interpretation:** The sensitivity, specificity, speed, window of clinical intervention, and ease of operation offered by the CRISPR-BP34 support its use as a point-of-care diagnostic for melioidosis.

**Funding:** Chiang Mai University Thailand and Wellcome Trust UK

**Research in context:** *Evidence before this study:* Melioidosis is an often-severe infectious disease caused by the bacterium *Burkholderia pseudomallei*. It is estimated to affect 165,000 individuals annually worldwide, of which 89,000 cases are fatal. The disease diagnosis is challenging due to diverse clinical presentations, low awareness, limited diagnostic options, or even a lack of diagnostic tests. A PubMed search conducted from the database inception to 6 May 2023, using the terms “melioidosis” AND “diagnosis test,” yielded 207 results, 40 of which presented clinical evaluations of rapid melioidosis diagnostic tests. Antigen-based diagnostic tests, which detect the presence of *B. pseudomallei*, reported high specificity (median = 98·6%, IQR 94·0 - 100·0), but low sensitivity (median = 57·1%, IQR = 44·3 - 82·5). The test sensitivity suffers from the often-low concentration of the bacterial antigens in patients’ samples, which can vary by specimen type and stage of infection. Antibody-based diagnostic tests that detect host antibodies against *B. pseudomallei* typically exhibit satisfactory specificity (median = 94·5%, IQR = 88·6 - 96·2) but poor sensitivity (median = 80·2%, IQR = 71·0 - 88·1). These tests are often impacted by variations in antibody responses to *B. pseudomallei* and the duration required for antibody production. Furthermore, standardisation remains challenging due to the influence of different serum titres on sensitivity and background of the tests. Likewise, quantitative PCR exhibits a high degree of specificity (median = 99·8%, IQR = 91·6-100·0), but an observed low sensitivity for melioidosis (median = 77·1%, IQR = 20·8-97·8), which is likely attributed to the genetic heterogeneity of *B. pseudomallei* genomes. Additionally, these studies consistently reported a demand for improved speed and ease of implementation in resource-limited settings where melioidosis is endemic. With the limitations of current diagnostic methods, a culture-confirmed approach with 60% sensitivity, 100% specificity, and a diagnosis time of 2-7 days still stands as the gold standard for melioidosis diagnosis.

*Added value of this study:* To date, no study has measured the impact of delayed diagnosis on melioidosis. We assessed the number of deaths occurring prior to culture-confirmed diagnosis (22·7%) and those after diagnosis but within 28 days post-admission (26·3%), highlighting the urgent need for prompt action. To address this, we developed the CRISPR-BP34 test, which utilises isothermal amplification of a nucleic acid target followed by site-specific detection using a CRISPR-Cas12a enzyme. We successfully implemented this assay in a resource-limited setting in northeast Thailand, where the disease prevalence is among the highest in the world. The assay achieved a diagnostic sensitivity and specificity of 93·0% and 96·8%, respectively, with a limit of detection ranging from 50-250 cfu/mL. Early diagnosis can be achieved within four hours of patient admission, which is significantly faster than the gold-standard test that typically takes several days. Moreover, the ultrasensitivity of the CRISPR-BP34 assay enabled the detection of low levels of *B. pseudomallei* in hemoculture bottles, which could be missed due to mixed infections, poor aseptic technique, or other causes, leading to undiagnosed melioidosis.

*Implications of all available evidence:* The CRISPR-BP34 assay holds great promise for the management and control of melioidosis. Its minimal setup and shallow learning curve make it well-suited for resource-limited settings. Additionally, its speed and high sensitivity enable early diagnosis and treatment, which are crucial for saving patients’ lives.

## INTRODUCTION

Melioidosis is a neglected tropical disease with a high case-fatality (10-50%) even when appropriately treated^1^. The global disease burden expressed in disability-adjusted life-years (DALYs) is 4·64 million, 99% of which is accounted for by years of life lost (YLL)^2^. High mortality may be explained by the disease disproportionately affecting rural populations in low-and middle-income countries (LMICs)^3^, many of which have poor socioeconomic conditions and often present to healthcare facilities in a terminal or critical phase of the disease^4^. Melioidosis is caused by *Burkholderia pseudomallei*, an environmental bacterium in soil and water across the tropical regions of Asia Pacific^5^, South^6^ and Southeast Asia^7^. However, the disease remains largely underreported due to its non-specific clinical manifestations that can “mimic” several other diseases. A lack of disease awareness in clinics and communities as well as the paucity of diagnostic facilities leads to missed or delayed diagnosis. With early diagnosis and appropriate treatment the mortality rate from melioidosis can be decreased to less than 10%^5^.

Clinical specimens from patients with suspected melioidosis are typically screened for the presence of *B. pseudomallei* using microbial culture, which has been the gold standard diagnostic method for the last three decades. This method is imperfect, with a specificity of 100% but a sensitivity of 60%^8^. *B. pseudomallei* grows more slowly in the laboratory compared with other pathogens^9^, which can lead to over-growth by other bacteria or fungi present in the sample as part of a mixed infection and/or contamination. This can result in failure to detect *B. pseudomallei*. If cultured successfully, *B. pseudomallei* colonies can be easily misinterpreted as environmental contaminants, and correct identification requires an expert microbiologist. Moreover, a combined time to grow and identify *B. pseudomallei* may take up to 7 days, which inevitably delays the disease diagnosis^8^. Culture-free antigen-based and nucleic acid-based tests such as a lateral flow immunoassay (LFI)^10^, an immunofluorescence assay (IFA)^11^, polymerase chain reaction (PCR)^12–14^, or 16S rRNA sequencing^15^ have been developed for the diagnosis of melioidosis. However, the heterogeneity in bacterial concentrations^16, 17^ across clinical specimens leads to a limited sensitivity of 58·2% (95% CI, 34·1 – 78·9%) for LFI^18^ and 63·8% (45·6 – 78·7%) for IFA^18^; whilst tests that offer higher sensitivity require thermal cyclers or sequencing machines^15^, which are rarely available in rural settings.

An improved sensitive and specific detection of *B. pseudomallei* DNA in a range of sample types using field-deployable equipment has the potential to improve the diagnosis of melioidosis. This can be achieved by first amplifying DNA of the pathogen using recombinase polymerase amplification (RPA), followed by sequence-specific recognition of clustered regularly interspaced short palindromic repeats (CRISPR)-Cas12a endoribonuclease at the DNA target. This approach has been applied to other bacterial pathogens including *Mycobacterium tuberculosis*^19^ and has been demonstrated to improve diagnosis and treatment responses. We previously described a robust CRISPR-based detection of genomic DNA of *B. pseudomallei in vitro*^20^. Here, we addressed the issues surrounding delayed diagnosis of melioidosis, established a diagnostic protocol for our recently developed CRISPR-Cas12a system (here-after termed CRISPR-BP34)^20^, and determined its diagnostic sensitivity and specificity. Our results showed higher sensitivity and shortened diagnostic time of the CRISPR-BP34 detection compared with the gold standard culture.

## METHODS

### Study design and patients

Two related studies were conducted and reported here. Study 1 quantified the time taken to diagnose melioidosis based on culture and patient outcome, and Study 2 evaluated the diagnostic performance of the CRISPR-BP34 assay (Figure 1).

**Figure 1:**
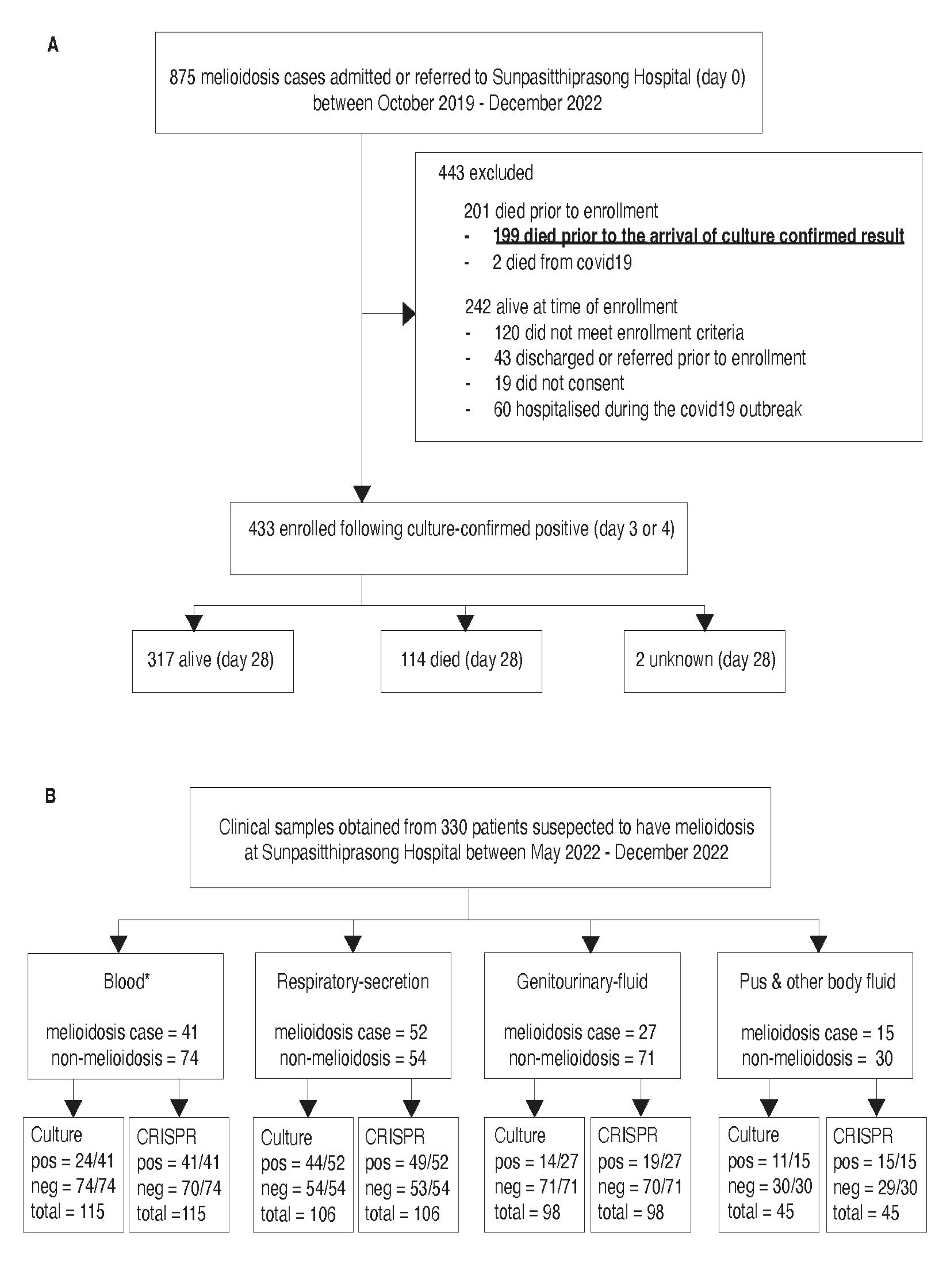
Study design: **A** summarises the mortality of melioidosis patients who were admitted or referred to Sunpasitthiprasong Hospital between October 2019 and December 2022. All cases were identified by culture-confirmed diagnosis, which commonly arrived on day 4. A substantial number of patients died prior to the arrival of the gold-standard test result. **B** presents our case-control study to evaluate the use of CRISPR-BP34 as an alternative rapid diagnostic test. Here, specimens were frozen upon arrival to enable subsequent screening with the CRISPR-BP34 test and head-to-head comparison with culture method.

Study 1 was a prospective observational cohort study of patients with culture-confirmed melioidosis at Sunpasitthiprasong Hospital, a tertiary care hospital in Ubon Ratchathani, Thailand between October 2019 and December 2022 (Figure 1A, appendix p 3). Study objectives were to assess the disease prevalence, the time taken to reach a diagnosis of melioidosis, the empirical antibiotics prescribed prior to diagnostic confirmation, and patient survival during 28- day follow-up from the admission date. Routine clinical practice was to obtain blood, urine, respiratory secretions and fluid (sputum, tracheal suction, pleural fluid), and other body fluid and tissue as available (pus, limb tissue, and synovial fluid) for culture from patients with suspected melioidosis. All samples were collected according to WHO guidelines^21^. The standard culture methods used for each specimen type are outlined in the appendix (p 5). All patients with culture-confirmed melioidosis were identified through the hospital computer. The length of time between sampling and culture result was recorded for each patient. Cases who remained alive when the culture result become available were visited to obtain written consent to collect clinical information. 875 culture-confirmed melioidosis cases were identified, of whom 433 were alive at the time of culture confirmation and gave written consent to obtain and use clinical information on antibiotic treatment and 28-day outcome (Figure 2). The study received ethical approval from the Sunpasitthiprasong Hospital Ethical Review Board (015/62C) and the Oxford Tropical Research Ethics Committee (OxTREC, 25-19). This study is registered with Thai Clinical Trial Registry (TCTR20190322003).

**Figure 2:**
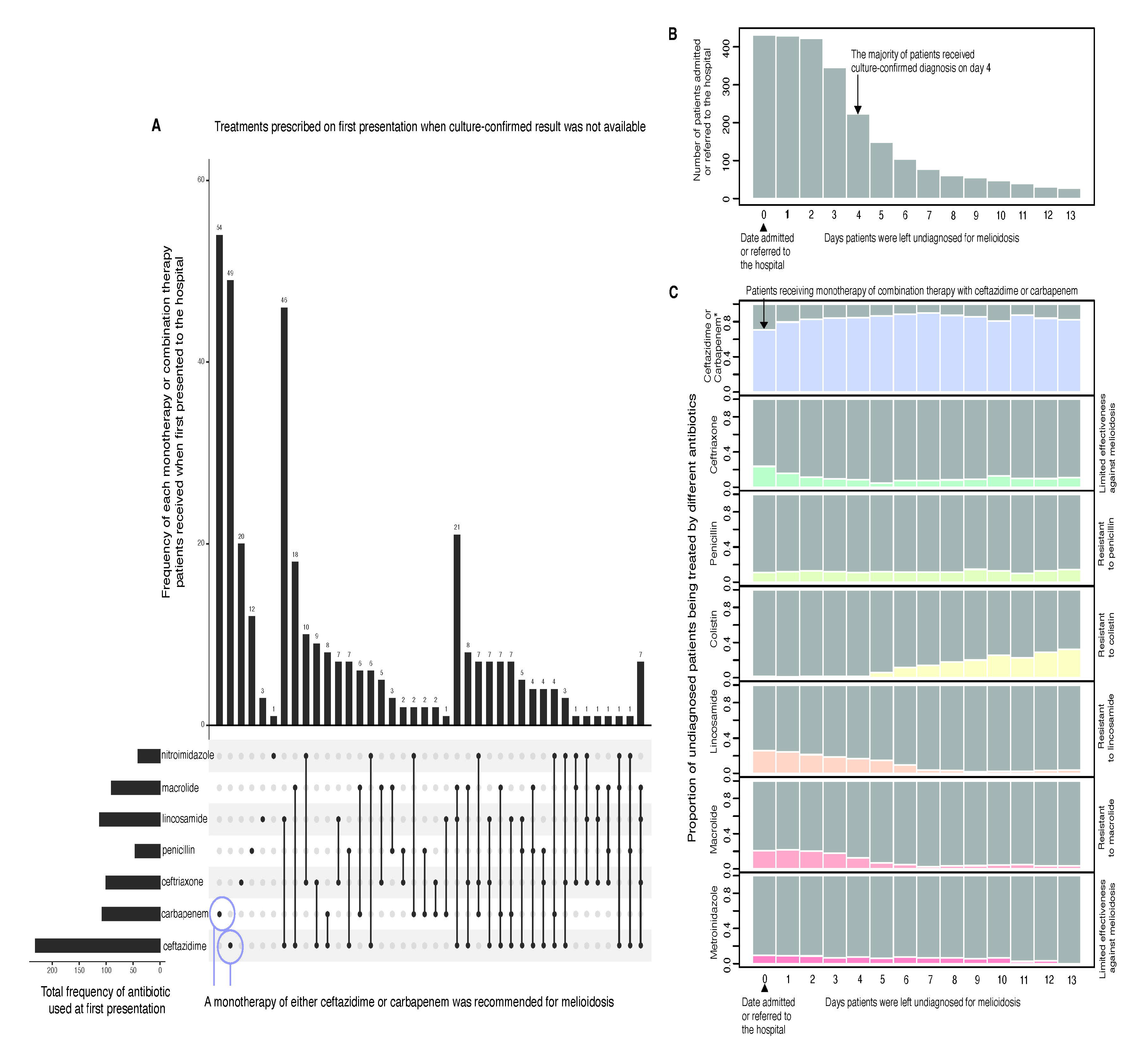
Burden of delayed diagnosis on melioidosis treatments and outcomes: **A** an UpSet plot summarises the antibiotic prescription when patients were first presented to Sunpasitthiprasong either as a monotherapy or combination therapy. **B** a histogram presents the number of patients who remained undiagnosed at a daily interval following their first presentation or referral to Sunpasitthiprasong Hospital during October 2019 to December 2022. Black arrow denotes the date when a culture-confirmed melioidosis result was available for most patients. **C** summarises choices of antibiotics being prescribed to patients who remained undiagnosed for melioidosis each day following their admission or referral date. Prescription of ceftazidime or carbapenem drugs either as monotherapy or combination therapy are highlighted in blue, with black arrow marking first day presentation.

Study 2 was a diagnostic evaluation of the sensitivity and specificity of a CRISPR-BP34 prototype assay^20^ conducted at Sunpasitthiprasong Hospital between May 2022 and December 2022 (Figure 1B). Minimal sample size was determined using the formula n = z^2^ p(1-p) /d^2^ where "z” is the 95% confidence interval at 1·96; “p” is the prevalence at 0·5; and “d” represents the margin of error at 0·1. At least 96 melioidosis and 96 non-melioidosis patients were required for the CRISPR-BP34 diagnostic test. Specimens were collected from patients with suspected melioidosis (appendix p 4) at the time of hospital presentation for culture identification. 1 mL of leftover sample from the culture was obtained from the hospital microbiology laboratory and stored at −20 °C for CRISPR-BP34 screening. Once the culture results arrived (appendix p 5), a head-to-head CRISPR-BP34 assay (appendix pp 7-9 & 10-16) was performed. In total, the study examined 114 melioidosis and 216 non-melioidosis patients, which were sampled across four different specimen types: blood (41 and 74 samples), respiratory secretions and fluids (52 and 54 samples); urine (27 and 71 samples), and other body fluids and tissues (15 and 30 samples) from melioidosis and non-melioidosis patients, respectively. The study received ethical approval from the Ethical Review Board of Sunpasitthiprasong Hospital (029/65C) and received ethical exemption from Chiang Mai University (9190/2565).

### Determining the molecular sensitivity of CRISPR-BP34 assay *in vitro*

To determine the limit of detection of the CRISPR-BP34 assay, we conducted *in vitro* experiments by inoculating genetically modified *Escherichia coli* that harbors a target DNA of the CRISPR-BP34 in its genome (appendix p 6) into blood and urine samples from a single healthy donor at different concentration ranging from 0, 10, 50, 100, 250, 500, 2500 and 5000 cfu/mL. Two to five biological replicates were performed for each experiment. CRISPR-BP34 detection was performed on blood as described in the appendix (pp 7-8). Blood and urine were selected to represent the most common types of clinical specimens processed from patients with suspected melioidosis. The use of the modified *E. coli* as a surrogate for *B. pseudomallei* was to circumvent laboratory safety requirement.

### Clinical evaluation of CRISPR-BP34 assay

Different types of clinical sample require specific sample preparation due to the variable amounts of target bacterial cells^16^, host cells and inhibitors present in each sample. Further details of how the assay was performed on each sample type is described in the appendix (pp 7- 8). For each clinical sample, human cells were first depleted using a simple buffer system to selectively lysed human cells, leaving a pellet of bacterial cells. Bacterial genomic DNA was then extracted from the pellet using either hot alkaline lysis or a spin column, depending on the pellet size. *B. pseudomallei* DNA was amplified in an RPA reaction, and the resulting amplicons were added into a 50-μL CRISPR reaction, comprising of CRISPR RNA (crBP34)^20^, LbCas12a protein and FAM-biotin probes. This reaction was incubated at 37 °C for 60 minutes, after which a HybriDetect lateral flow dipstick (TwistDx, UK) was directly immersed into the reaction and allowed to develop for 5 minutes before reading by eye. A positive result was defined as the appearance of an upper band (anti-IgG) on the dipstick. The assay was performed in a batch of 10 samples with each batch comprising of culture positive and negative samples to avoid batch effect. For all batches, *B. pseudomallei* positive sample and distilled water were used as positive and negative controls, respectively. The sample-to-result time was recorded. The CRISPR-BP34 results were interpreted by three different readers who were blinded to the patient disease status and culture results.

Not every sample cultured from a melioidosis patient yields positive result, a scenario driven by different local concentration of *B. pseudomallei*. Discordant results between culture and the CRISPR assay were tested by quantitative PCR (qPCR) using three primer sets listed in the appendix (pp 9, 19-20). Given a large discrepancy in reported diagnostic sensitivity of PCR primers^18^ and a high sequence diversity of *B. pseudomallei*, the use of primer combinations ensured an increased coverage of the detection. The qPCR cycle threshold (ct) values were recorded and used as a proxy for bacterial loads.

### Statistical analysis

All numerical data was summarised using medians, interquartile ranges (IQR), and proportion. For comparison with non-parametric distribution, Wilcoxon signed-rank tests were used with Benjamini-Hochberg correction for multiple testing. The culture and CRISPR-BP34 results from the same specimen were compared against the patient’s disease status. Sensitivity and specificity were defined as TP/ (TP + FN) and TN/ (TN + FP), respectively. “TP” and “FN” represent the number of cases that tested positive and negative in the disease group, respectively. “TN” and “FP” are the number of cases that tested negative and positive in the non-disease group, respectively. The sensitivity and specificity were separately calculated for culture and CRISPR by sample type (grouped into four categories: blood; urine; respiratory secretion; and pus, tissue and other body fluid), and by total specimens. For a patient who had multiple samples taken, only the earliest sample was included for sensitivity and specificity calculations. The exact 95% confidence interval was estimated using Clopper-Pearson Binomial assumption^30^.

## RESULTS

### Time to diagnosis, empirical antibiotic treatment, and outcomes

Of 875 culture-confirmed melioidosis patients admitted or referred to Sunpasitthiprasong Hospital during October 2019 to December 2022, 199 died before the culture results arrived (Figure 1A). Among 433 patients who survived to culture results and consented to participate in our study, 114 patients died within 28 days after first presentation to Sunpasitthiprasong hospital. The minimum fatality of 35·8% based on the combined data is consistent with 35% mortality reported in Thailand^22^. A relatively high fatality reported in this setting could be attributed to socioeconomic disparity^2^ which impacted mostly agricultural workers (272 [62·8%] of 433 patients) who lived on average 65 km (IQR 40 – 100 km) from the culture-confirmed diagnosis facility (Table 1). Melioidosis patients displayed diverse clinical manifestations with persistent fever being the most common symptom (Table 1, appendix p 18. Patients experienced symptoms for a median of seven days (IQR 3 −14 days) before seeking medical care at the local or central health-care centres, which subsequently referred them to Sunpasitthiprasong hospital within one day (IQR 0 – 4 days). Multiple samples were collected from patients for culture diagnosis upon first admitted or referred, and during their stay at Sunpasitthiprasong hospital as clinically indicated. The median duration between the first sample collection and the first positive culture result was four days (IQR 3 - 5 days), at which point melioidosis diagnosis was confirmed. Notably, the time from symptom onset to diagnosis averaged 16 days (IQR 9 – 27 days), although this differed significantly between patients who died (12 days, IQR 7 – 19 days) and those who survived (18 days, IQR 10 – 31 days, Wilcoxon test p-value = 5·62 x 10^-5^), underscoring the need for prompt clinical intervention to avert fatal outcomes.

**Table 1.**
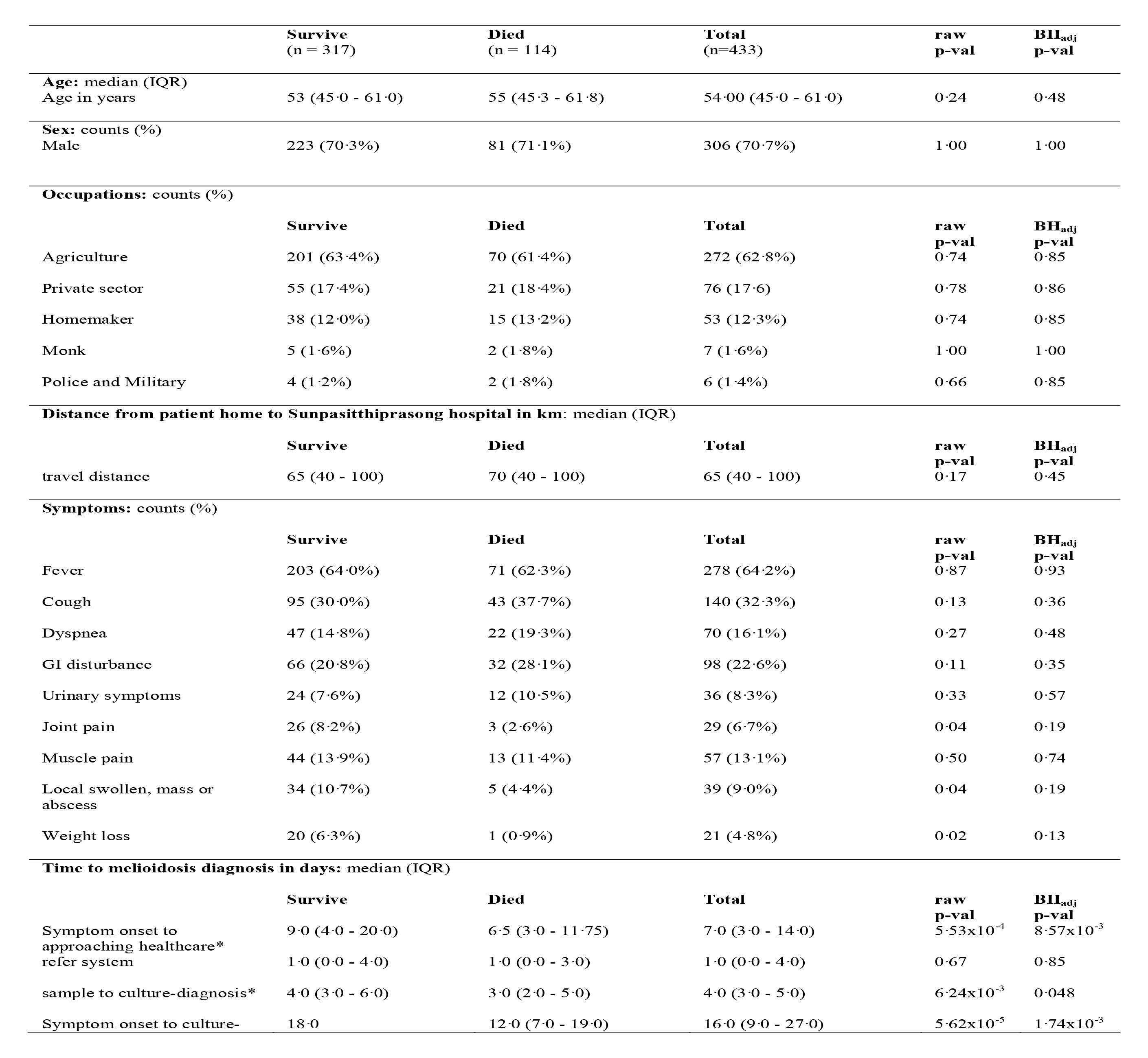
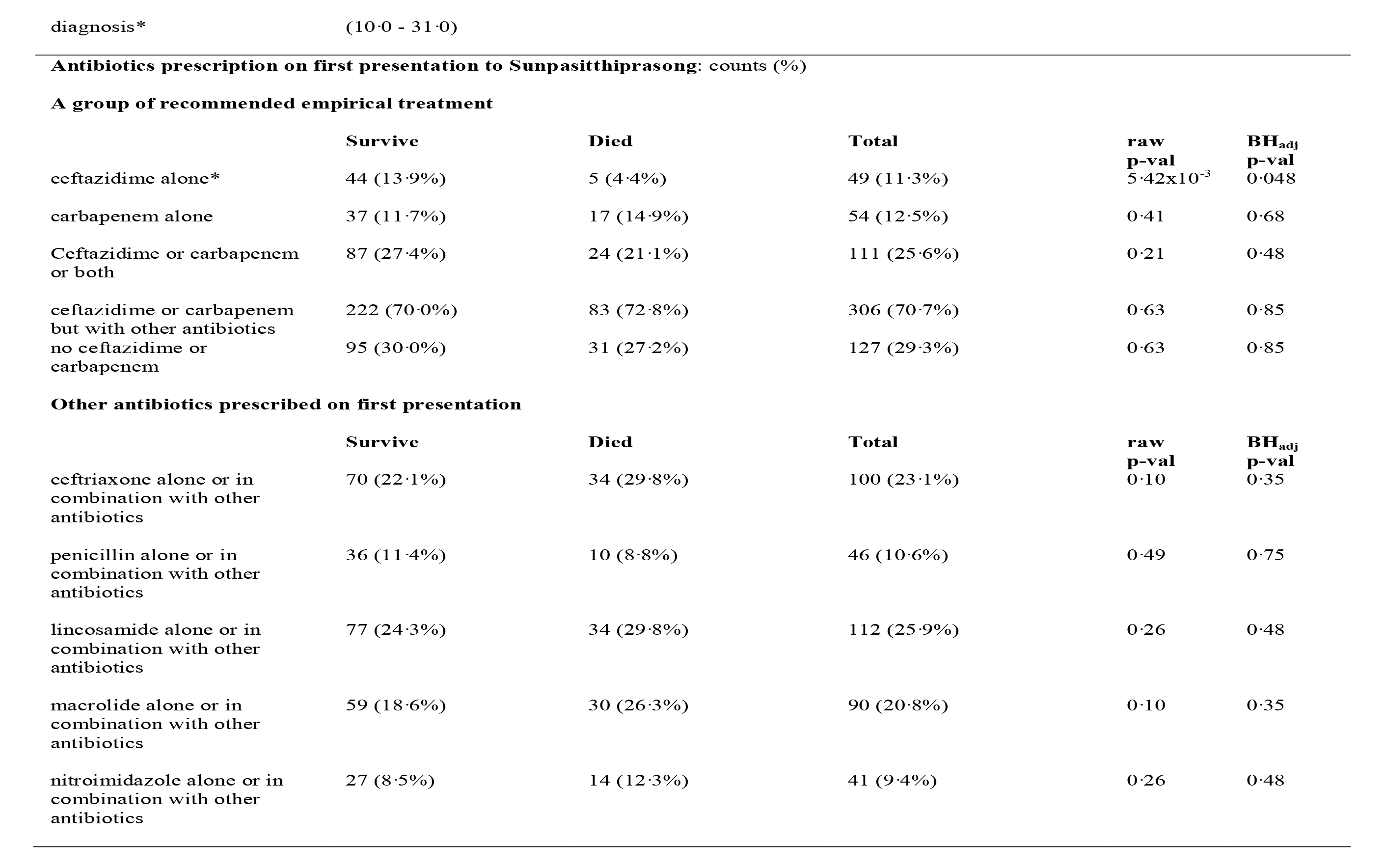
Demographic of melioidosis patients admitted or referred to Sunpasitthiprasong hospital and survived to culture results during October 2019 to December 2022. Significant terms following Benjamini-Hochberg correction for multiple testing were marked with asterisks

In Thailand, patients suspected of having melioidosis are recommended to receive empiric treatment with intravenous ceftazidime or a carbapenem antibiotics, such as meropenem or imipenem, for initial intensive monotherapy^23^. Antibiotic administration data was available for 433 patients of which 49 (11·3%) and 54 (12·5%) respectively received ceftazidime or a carbapenem monotherapy upon their first presentation to Sunpasitthiprasong hospital (Figure 2A, Table 1). The remainder received other treatments including a monotherapy or combinations of antibiotics known to be ineffective in treating melioidosis such as lincosamides^24^, macrolides^24, 25^, penicillin^25^ and first-and second-generation cephalosporin^24, 26^, and ineffective third-generation cephalosporins such as ceftriaxone^27^; an inevitable practice to cover the broad spectrum of infection when the causative agents were unknown (Figure 2B & C). A lower mortality (5 [10·2%] of 49) was observed in patients who received ceftazidime monotherapy than in patients who received other types of treatments (109 [28·5%] of 382; Fisher’s exact p-value 5·42 x 10^-3^, Table 1). To increase the use of ceftazidime monotherapy at first presentation from 11·3% to 100%, a more rapid diagnosis will be necessary.

### Clinical evaluation of the CRISPR-BP34 assay

We developed the CRISPR-BP34 assay to detect *B. pseudomallei* DNA, which demonstrated high sensitivity and specificity *in vitro*^20^. We tested the limit of detection of the CRISPR-BP34 by spiking blood and urine samples and found it to be 250 cfu/mL and 50 cfu/mL, respectively (Figure 3A & B). To determine whether CRISPR-BP34 test is sufficiently sensitive to detect the bacterium across various clinical samples, we enumerated the numbers of *B. pseudomallei* recovered from different specimen types using both new experiments and previous datasets^16, 17^ (Figure 3C). Our results showed that the number of *B. pseudomallei* detected in most common specimens such as urine (median = 2·6 x 10^4^ cfu/mL, sputum (median = 8·8 x 10^7^ cfu/mL), pus and other body fluids (median = 5·4 x 10^7^ cfu/mL) were greater than the CRISPR-BP34 limit of detection. Since direct blood samples had a lower *B. pseudomallei* concentration (median = 1·5 cfu/mL) than the limit of detection, we substituted direct blood samples with hemoculture positive samples (median = 7·3 x 10^7^ cfu/mL) to ensure sufficient bacterial concentration. Hemoculture positive samples are blood samples that have been incubated in a culture medium to allow the bacterium to grow, but at this point, the bacterium’s identity remained unknown (Figure 3D).

**Figure 3:**
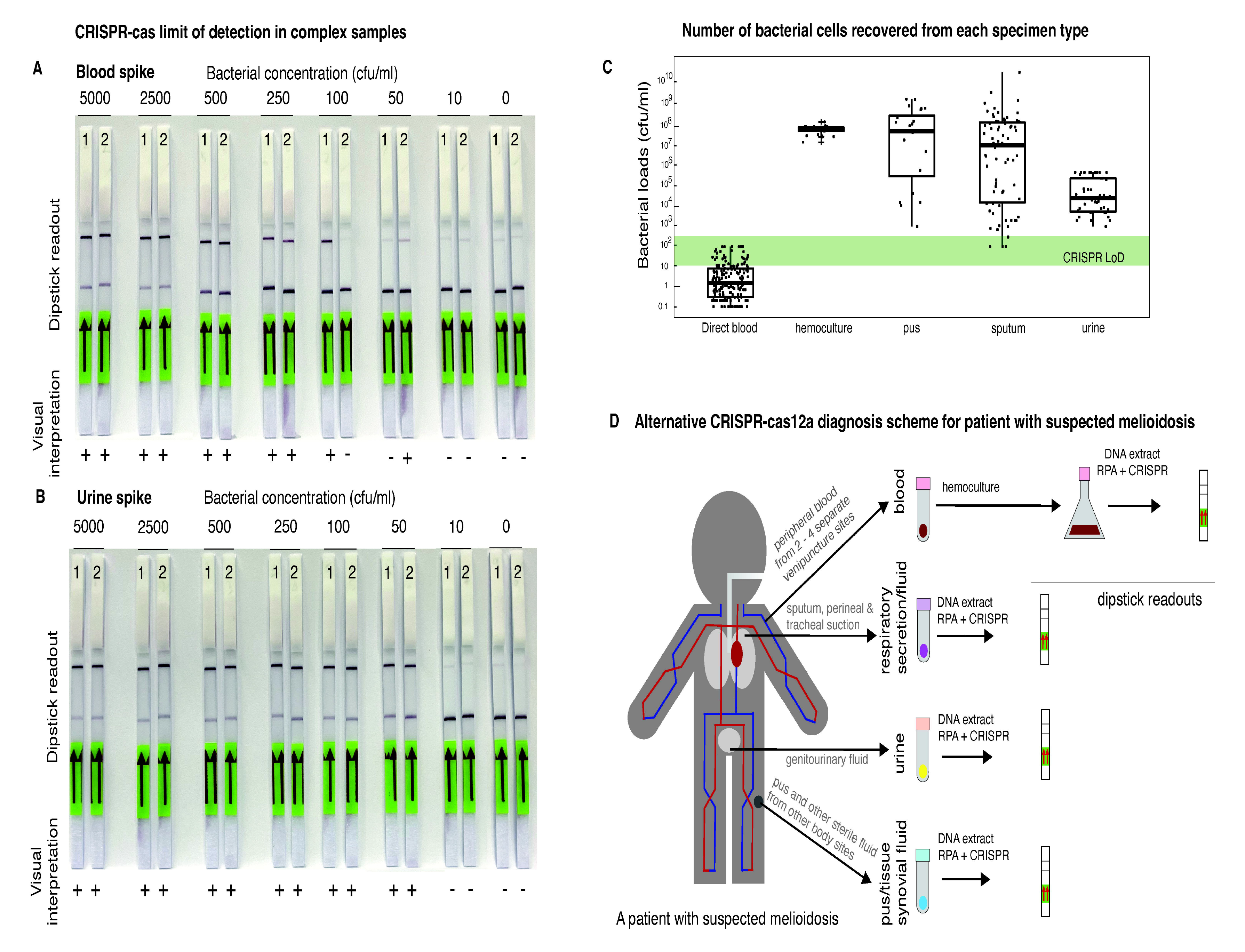
Limit of detection of the CRISPR-BP34 assay and its proposed diagnostic pipeline. **A** and **B** present the limit of the CRISPR-BP34 test detection in blood and urine samples which are the two most common specimens, respectively. The dipsticks were shown as two biological replicates with LOD of urine (50 cfu/mL) and blood (250 cfu/mL). **C** summarises the number of *Burkholderia pseudomallei* that can be recovered from different type of specimens. Green bar indicates the range of limit of detection of the CRISPR test. **D** presents an overview of CRISPR-BP34 diagnosis based on clinical samples including blood, genitourinary fluid, respiratory secretion, pus and other body fluid that were routinely collected from patients suspected of melioidosis. Bacterial pathogen in blood is typically enriched for growth to the detectable threshold (250 cfu/mL) through hemoculture process.

We tested the hypothesis that CRISPR-BP34 assay would be more sensitive and faster than culture-confirmed approach in clinical samples collected from 114 melioidosis and 216 non-melioidosis patients. Of these, 20 melioidosis and 12 non-melioidosis patients had samples collected across multiple specimen types (Figure 4A), while 94 melioidosis and 204 non-melioidosis patients had a single sample type collected (Figure 4B). Using first sample available from each patient, we estimated the overall diagnostic sensitivity and specificity of both methods. Our findings showed an overall sensitivity of CRISPR-BP34 of 93·0% (106 of 114 samples, 95% CI 86·6 – 96·9), higher than the sensitivity of culture at 66·7% (76 of 114 samples, 95% CI 57·2 - 75·2) (Figure 4C, Table 2). The overall specificity of the CRISPR-BP34 was 96·8% (209 of 216 samples, 95% CI 93·4 - 98·7), compared to 100% (216 of 216 samples, 95% CI 98·3 - 100) for culture (Figure 4D, Table 2). Sensitivity of CRISPR-BP34 versus culture for individual sample types was generally higher for CRISPR-BP34, as follows: hemoculture at 100% (41 of 41 samples) vs 58·5% (24 of 41 samples); urine at 70·4% (19 of 27 samples) vs 51·9% (14 of 27 samples); respiratory secretions and fluids at 94·2% (49 of 52 samples) vs 84·6% (44 of 52 samples); and other body fluids and tissue at 100% (15 of 15 samples) vs 73·3% (11 of 15 samples) in CRISPR-BP34 (Figure 4C and Table 2). Specificity of CRISPR-BP34 versus culture for individual sample types was equivalent or slightly lower for CRISPR-BP34, as follows: hemoculture at 94·6% (70 of 74 samples) vs 100% (74 of 74 samples); urine at 98·6% (70 of 71 samples) vs 100% (71 of 71 samples); respiratory secretions and fluids at 98·2% (53 of 54 samples) vs 100% (54 of 54 samples); and other body fluid and tissue at 98·67% (29 of 30 samples) vs 100% (30 of 30 samples) in CRISPR-BP34 and culture assay, respectively (Figure 4D and Table 2).

**Figure 4.**
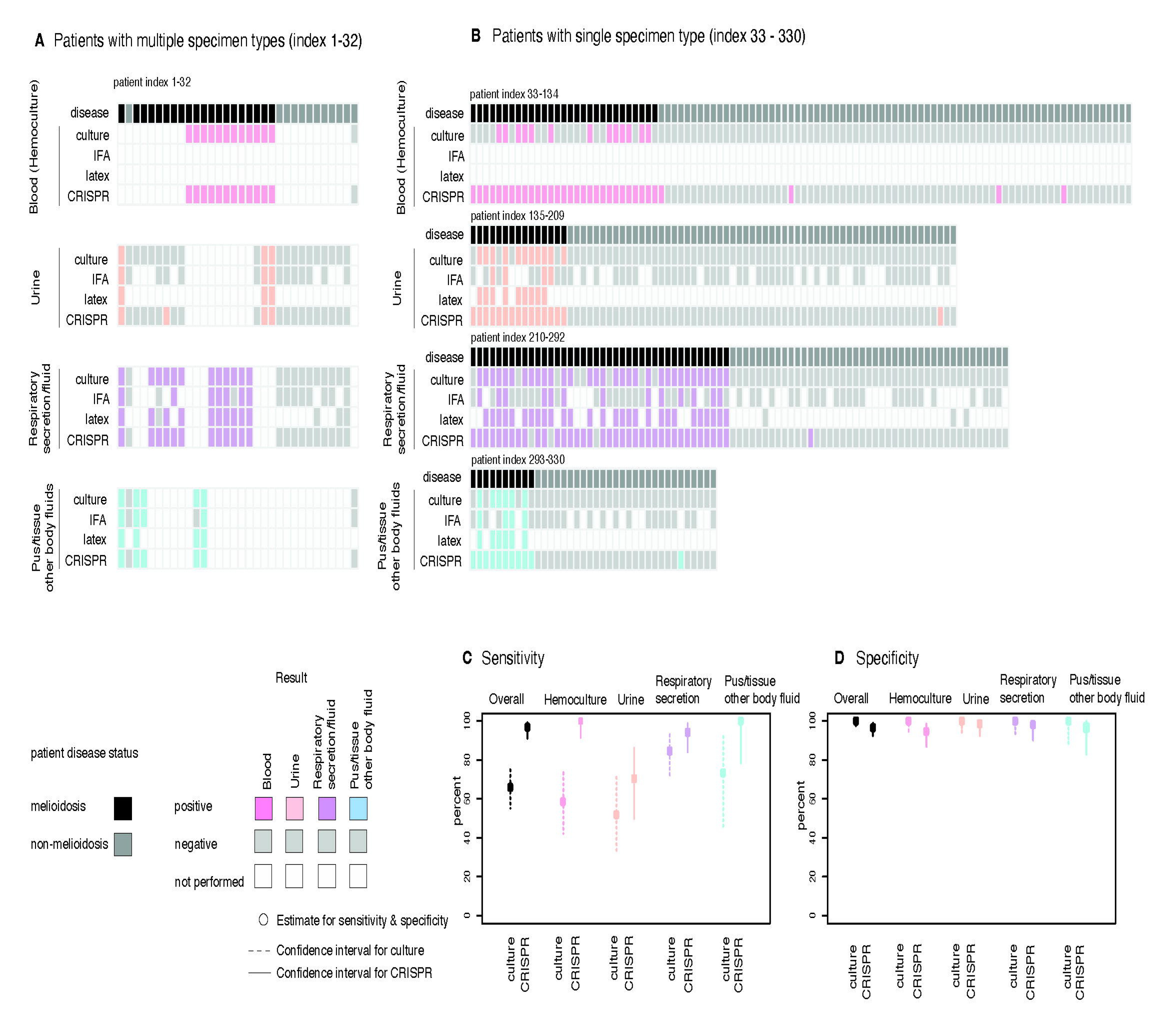
Sensitivity and specificity of the culture and the CRISPR approaches. **A** present combined panels of diagnostic results from 32 patients with multiple specimen types. Top to bottom panels summarise results from hemoculture, genitourinary fluid, respiratory secretion, and pus or other body fluids, respectively. For patients with multiple samples per each sample type, only the first sample was presented. Rows in each panel display results from different diagnostic tests including the gold-standard culture, IFA, latex agglutination, and the CRISPR-BP34 test (top to bottom). Each column corresponds to data from each patient with disease status marked as melioidosis (black) or non-melioidosis (grey). **B** summarises findings from 298 patients with single specimen type with results from hemoculture, genitourinary fluid, respiratory secretion and pus or other body fluids presented in the same order as in (**A**). **C** and **D** compare the sensitivity and specificity of the gold-standard culture and the CRISPR-BP34 approach, respectively. Dots indicate the actual sensitivity or specificity values while the 95% confidence interval are represented by dotted line (culture) and solid line (CRISPR-BP34).

**Table 2.** Clinical sensitivity and specificity of culture and CRISPR-based methods.

| Sample types | Disease status |  | Culture-based approach |  | CRISPR-BP34 |  |
| --- | --- | --- | --- | --- | --- | --- |
|  | Melioidosis cases | Non-melioidosis controls | Sensitivity<br>(Percent, 95% CI) | Specificity<br>(Percent, 95% CI) | Sensitivity<br>(Percent, 95% CI) | Specificity<br>(Percent, 95% CI) |
| Overall | 114 | 216 | 76 of 114<br>(66.7%,<br>CI 57.2 – 75.2%) | 216 of 216<br>(100%,<br>CI 98.3 – 100.0%) | 106 of 114<br>(93.0%,<br>CI 86.6 – 96.9%) | 209 of 216<br>(96.8%,<br>CI 93.4 – 98.7%) |
| Hemoculture | 41 | 74 | 24 of 41<br>(58.5%,<br>CI 42.1 – 73.7%) | 74 of 74<br>(100%,<br>CI 95.1 – 100.0%) | 41 of 41<br>(100.0%,<br>CI 91.40 – 100.0%) | 70 of 74<br>(94.6%, CI 86.7 –<br>98.5%) |
| Genitourinary fluid | 27 | 71 | 14 of 27<br>(51.9%,<br>CI 32.0 – 73.3%) | 71 of 71<br>(100%,<br>CI 94.9 – 100.0%) | 19 of 27<br>(70.4%,<br>CI 49.8 – 86.3%) | 70 of 71<br>(98.6%,<br>CI 92.4 – 100%) |
| Respiratory secretion | 52 | 54 | 44 of 52<br>(84.6%,<br>CI 71.9 – 93.1%) | 54 of 54<br>(100%,<br>CI 93.4 – 100.0%) | 49 of 52<br>(94.2%,<br>CI 84.1-98.8%) | 53 of 54<br>(98.2%,<br>CI 90.1-99.6%) |
| Pus, tissue and other body fluids | 15 | 30 | 11 of 15<br>(73.3%,<br>CI 44.9-92.2%) | 30 of 30<br>(100%,<br>CI 88.4 -100.0%) | 15 of 15<br>(100%,<br>CI 78.2 – 100.0%) | 29 of 30<br>(96.67%,<br>CI 82.8-99.9%) |

In addition, the CRISPR-BP34 assay provided faster results and significantly reduced the turnaround time for all sample types compared to culture approach (Figure 5A, Wilcoxon test p-value < 2·2 x 10^-16^ for all sample types). For culture positive samples, the median sample-to-result time was 2·5 days (IQR 1·8 – 3·3 days) for hemoculture, and 3·9 days (IQR 3·7 – 4·1 days) for urine, respiratory secretion and fluids, as well as other body fluids and tissues. In contrast, the average sample-to-result time for the positive CRISPR-BP34 assay was 1·1 days (IQR 0·7 – 1·5 days) for hemoculture, 2·3 hours (IQR 2·3 – 2·4 hours) for urine, and 3·3 hours (IQR 3·1 – 3·4 hours) for respiratory secretion, and fluids, as well as other body fluids and tissues.

**Figure 5.**
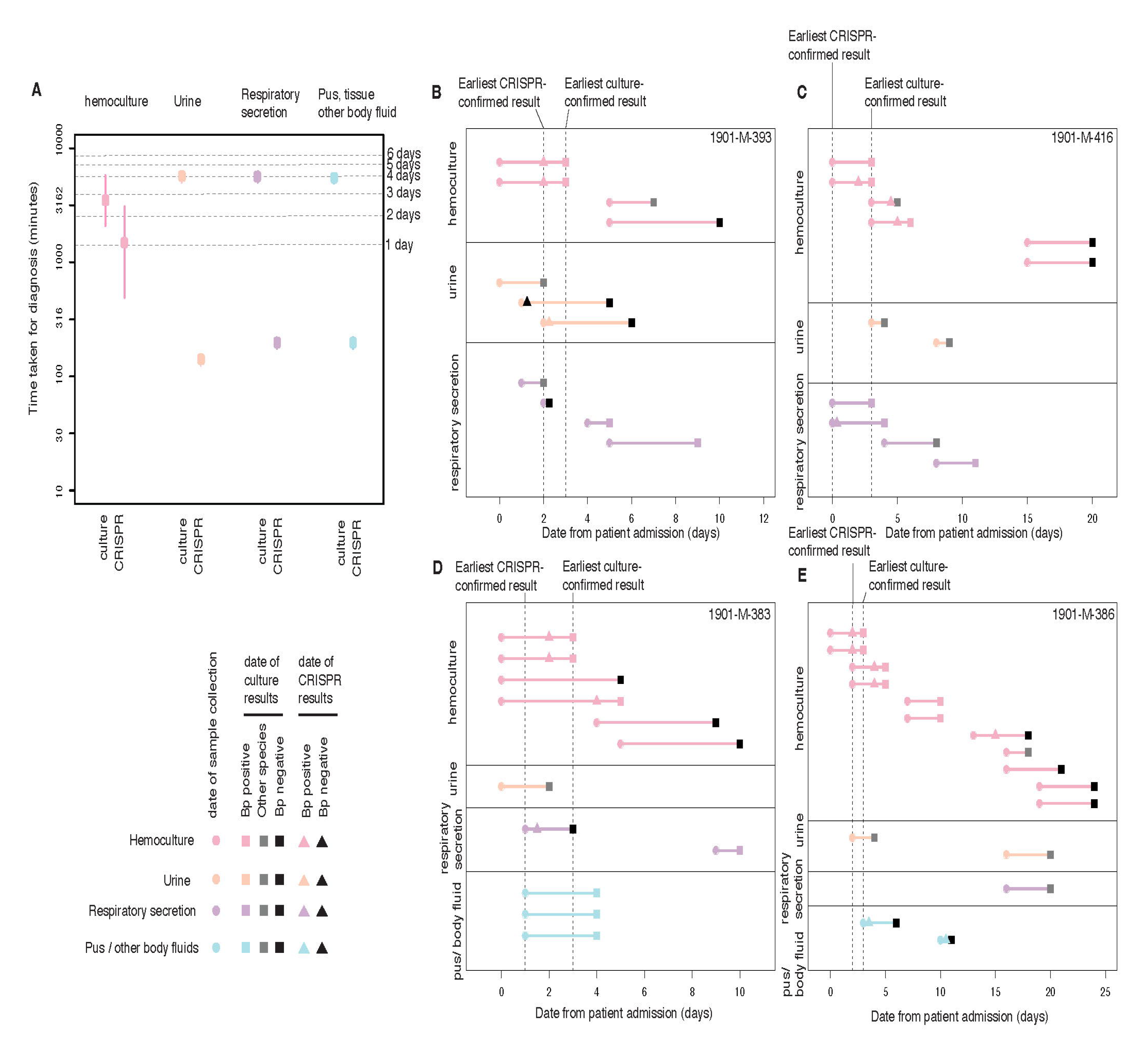
Time taken for the culture and the CRISPR-BP34 approaches, and case examples. **A** illustrates the diagnostic time required for the culture and the CRISPR approaches on the log 10 scale. **B** to **E** highlighted selected cases (4 of 20 melioidosis cases) where both multiple sample types and multiple specimens per sample type were available (vertical axis). The horizontal axis represents time in days from the date of admission or referral to Sunpasitthiprasong hospital. Circles mark the dates when samples were taken, while rectangles and triangles present the time when the culture-confirmed results and the CRISPR-BP34 results arrived, respectively. For both culture-and CRISPR-BP34 approaches; black denotes negative result, grey highlights detection of other bacterial species than *B. pseudomallei*, while pink (hemoculture), orange (urine), purple (respiratory secretion) and blue (pus, tissue, and other body fluids) present *B. pseudomallei* positive result from each sample type.

### Potential use case

20 of 114 melioidosis patients had multiple sample types, as well as multiple numbers of samples per type collected over time (Figure 4A, Figure 5), enabling investigation of how each patient was diagnosed and treated in the real world. Early specimens were collected on the first or within a few days of admission or referral to aid disease diagnosis, while late specimens were taken at 3-, 5- or 7-days intervals following antibiotic prescription to assess response to the treatment. A proportion of early specimens taken from patients who were later confirmed to have melioidosis were culture negative. This could be either attributed to the concentration of *B. pseudomallei* being lower than the culture limit of detection, or contamination of the samples with other fast-growing bacterial species (Figure 5B to D). One striking example was a hemoculture bottle from a melioidosis case presented in Figure 5C, which was contaminated with coagulase-negative staphylococci, a skin commensal. The CRISPR-BP34 assay, however, reported *B. pseudomallei* positive from this contaminated blood bottle, consistent with subsequent qPCR experiments confirming the presence of *B. pseudomallei* DNA and with patient’s final diagnosis. We observed a high cycle threshold (ct) score of *B. pseudomallei* in the contaminated hemoculture bottle and other contaminated samples (median = 32·3, IQR 30·5 - 35·9), compared to average ct values detected in *B. pseudomallei* positive hemoculture (median = 14·0, IQR 13·4 - 14·9, appendix p 17). This likely indicates that the *B. pseudomallei* population was outcompeted by contaminant species. Cross-contamination incidents like this are not uncommon in laboratories with limited resources such as in rural Thailand which could result in an underestimation of the true incidence of melioidosis^28^. Regardless, for all cases being followed, CRISPR-BP34 provided earlier identification of *B. pseudomallei* than the culture method (Figure 5B to D).

## DISCUSSION

The first part of our study documented the time taken from first melioidosis symptom-to-diagnosis, the empirical treatment patients received during diagnosis uncertainty, and death that occurred prior or after the arrival of culture-confirmed diagnosis result. We noted that many severely ill patients died before we could reach them, thereby introducing survival bias in our study. Nevertheless, our findings echo the problems of delayed disease diagnosis, and imperfect treatment during diagnosis uncertainty, which may separately, or collectively contribute to death. To improve the appropriate and timeous initiation of melioidosis treatment, we developed an easy to implement point-of-care CRISPR-BP34 assay (appendix pp 7-16), tested the sample-type specific protocols (Figure 3D), and evaluated the test performance (Figure 4) in the second part of our study.

To our knowledge, a lower limit of detection at the range of 50 – 250 cfu/mL is the highest among reported melioidosis rapid diagnosis tests without requiring extensive equipment such as a qPCR machine or a UV microscope. For common clinical specimens with high bacterial loads (over 10^3^ cfu/mL) such as hemoculture samples, genitourinary fluids, respiratory secretions, and pus and other body fluids; CRISPR-BP34 could be used on DNA extracted from these direct samples. This resulted in high level of sensitivity of the CRISPR-BP34 test of 93·0% compared to 66·7% sensitivity for overall samples of culture approach (Figure 4C). Although CRISPR-BP34 could detect the presence of *B. pseudomallei* at the concentration as low as 50 cfu/mL, the miniscule volume of specimens utilised by the CRISPR approach means that the test can be skewed by inaccurate pipetting or handling errors. Thus, for direct blood samples, we recommend using CRISPR-BP34 on DNA extracted from hemoculture (enriched media) instead of direct blood samples to maintain high sensitivity.

We also observed a slight drop in the CRISPR test specificity being 96·8% compared to 100% specificity of culture approach across all sample types (Figure 4D). Some of the “false” positives could be “true” but “missed” diagnosis cases as result of current imperfect diagnosis techniques including culture-confirmed approach^8^ and qPCR^18^ with suboptimal primers. Some “false” positives may arise when high copy numbers of genetic materials or RPA amplicons were mixed or handled in a confined bench setting, which is sometimes unavoidable in crowded space of resource-limited laboratories. Alternatively, the CRISPR-BP34 complex may mistakenly target other DNA sequences, resulting in false positives. However, the latter is likely mitigated by the double-layered specificity provided by RPA primers and CRISPR RNA; each of which were carefully designed using the genomic database of over 40,000 bacterial and human DNA^20^. To ensure robust test, suggestions to minimise the DNA cross-contamination which could generate false positives were fully documented in the appendix (p 9).

In summary, the CRISPR-BP34 test has exhibited higher sensitivity and comparable specificity to the gold standard culture-confirmed method, with more rapid detection capabilities across various types of clinical specimens. Our findings provide evidence to support the use of the CRISPR-BP34 test as a point-of-care diagnostic tool. Its implementation could lead to prompt initiation of lifesaving treatment (Figure 5B to E), with use cases discussed and positively received by Ministry of Public Health Thailand and regional health authorities^29^.

### Contributors

CChe and SPW conceived the study and secured funding. PC, SS, CU, HT, CChe and SPW, designed the study. SP, PB, KA, YD, AF, GW, PA, PK, VW, and CCho were involved in study implementation under CChe and SPW supervision. CChe and SPW did the analysis. CChe, SPW, PC, SJP, NPJD, NRT, and CU interpreted the data. CChe and SPW wrote the first draft. All authors read and approved the manuscript.

### Data sharing

Deidentified participant data that underlie the results reported in the article will be made available upon request. Proposals should be directed to the corresponding author. Proposals will be reviewed and approved by the sponsor on the basis of compliance with the informed consent and scientific merit.

## Data Availability

All data produced in the present study are available upon reasonable request to the authors

## Acknowledgements

SPW was funded by TSRI Fundamental Fund 2022, Chiang Mai University (grant number 65A104000009) and Faculty of Medicine Research Fund, Chiang Mai University (grant number MIC-2562-06372). CCho was funded by Wellcome International Master Fellowship (221418/Z/20/Z). CChe was funded by Wellcome International Intermediate Fellowship (216457/Z/19/Z) and Sanger International Fellowship. This research was funded in whole, or in part, by the Wellcome Trust. For the purpose of Open Access, the author has applied a CC BY public copyright licence to any Author Accepted Manuscript version arising from this submission.

