## Supplementary appendix for "Benchmarking CRISPR-BP34 for point-of-care melioidosis detection in LMIC: a molecular diagnostics study"

### SUPPLEMENTARY METHODS

#### Study 1 Protocol: a prospective observational cohort study of patients with culture-confirmed melioidosis

#### Study design, sites, and population

The objective of the study was to examine the clinical and genetic factors that influence disease outcomes (28-day mortality) in melioidosis. To achieve this, the study gathered clinical data including the time taken to reach melioidosis diagnosis, medical history, symptoms, and treatments received; bacterial samples, and individual blood samples from both melioidosis cases and control cases. The patients involved in this study were recruited from Sunpasitthiprasong Hospital in Ubon Ratchathani, where high incidence of melioidosis is observed. Additionally, the hospital houses the satellite unit of the Mahidol Oxford Tropical Medicine Research Unit (MORU) that facilitated the processing of clinical samples. Melioidosis patients were recruited from the Hospital Infectious department. The recruitment phase of the study lasted for three years (October 2019 - December 2022), covering three monsoon seasons when a rise in melioidosis cases was anticipated. Only the data concerning the host characteristics of the cohort was described in this paper.

#### Participant inclusion criteria

-Age ≥ 18 years
-Suspected cases of Melioidosis on admission or referral to the hospital
-Willingness to participate in the study and written, informed consent obtained from the patient or their relative.
-Resident in Northeast Thailand for at least the previous 2 years

#### Participant exclusion criteria

-Current TB or treatment of TB within the last 6 months
-Documented HIV infection, chemotherapy, or other immunosuppressant therapy in the last 12 months.
-Pregnancy
-Current covid19 (due to the recruitment during covid pandemic) *

#### Screening

**All** patients at Sunpasitthiprasong Hospital with a culture-confirmed diagnosis of suspected melioidosis were approached by a member of study staff. The patient were screened for eligibility. A written participant information sheet (PIS) and informed consent form (ICF) written in Thai language were presented to the participant or relative where they lack capacity detailing no less than: the exact nature of the study; the implications and constraints of the protocol; the known side effects and any risks involved in taking part. The participant or relative were allowed adequate time (minimum of ten minutes) to consider the information, and the opportunity to question the Investigator or other independent parties to decide whether they will participate in the study. Written informed consent were then be obtained by means of participant or relative dated signature and dated signature of the person who presented and obtained the informed consent. If the patient or relative is illiterate, a thumbprint was obtained. A copy of the signed Informed Consents was given to the participant or responsible relative. The original signed forms has been retained at the study site.

#### Ethics/protection of human subjects

The study received ethical approval from the Sunpasitthiprasong Hospital Ethical Review Board (015/62C) and the Oxford Tropical Research Ethics Committee (OxTREC 25-19). All research data sets arising from the study were pseudonymised. This allowed for eventualities such as feedback of individual data or withdrawal of consent. However, subject names or identities of participants will not be revealed.

**Symptoms of patients suspected to have melioidosis**

Patients presented with melioidosis display broad clinical manifestations. It is therefore clinically challenging to differentiate melioidosis from other infections. Patients suspected to have melioidosis presented with one or more of the following symptoms[1]. The frequency of symptoms observed in our collection was summarised in **Supplementary Figure S9**. Clinical specimens from these patients were thus tested for the presence of *B. pseudomallei.*

i. High and persistent fever: Fever is the most common symptom with melioidosis fever often lasting several days or weeks. The fever may start abruptly with chills and sweating. In melioidosis endemic areas such as our setting in Ubon Ratchathani, it can be challenging to discriminate melioidosis from other febrile illnesses based on fever alone.

ii. Respiratory symptoms: the severity of the symptom can vary depending on the location of infection and the bacterial loads. The common symptoms included persistent cough, chest pain, shortness of breath, hemoptysis or pneumonia.

iii. Skin lesions: the severity of the symptom can vary depending on the location. This can be manifested as small localised abscesses to large necrotic ulcers. Skin lesions can present with swollen areas on the skin and can be accompanied by lymphadenopathy.

iv. Gastrointestinal symptoms: the severity can vary depending on the extent of infection. The symptoms typically include abdominal pain, nausea and vomiting, diarrhea (can be watery or bloody), anorexia (which results in weight loss), gastrointestinal bleeding (which leads to anemia) and hepatomegaly.

v. Muscle and joint pain: Joint pain could either be localised to one area or affect multiple joints. The symptoms can be accompanied by stiffness, swelling and redness in the affected area. Similarly, muscle pain can be localised or generalised over the entire body, and can be accompanied by muscle weakness and fatigue.

vi. Neurological symptoms (reported in Australia with a small number of cases documented in Thailand): this included headache, seizures, altered mental status, cranial nerve dysfunction, motor weakness, sensory disturbances, and ataxia.

**Culture-confirmed assay used to screen patients**

**Blood sample**

As the amount of *B. pseudomallei* cells directly recovered from patient blood is low (1 CFU/mL), an enrichment process is required. Peripheral blood samples were aseptically drawn from venipuncture sites and transferred into BACTEC™ Plus Aerobic/F Culture Vials catalogue number 442192 (Becton Dickinson, Cockeysville, MD, USA). Blood culture bottles were incubated at 35 °C in the BACTEC 9120 instrument until a positive result or for a maximum of 5 days. As part of the laboratory routine, the causative pathogens were identified from positive bottles by subculturing the positive specimens on blood agar plates. The plates were incubated at 37 °C and inspected daily for the appearance of colonies, typically within 1-2 days (**Figure 3A**).

**Urine sample**

A urine sample was collected from a patient suspected of having melioidosis. As a part of routine practices, 1 µgL of an unprocessed urine sample (neat sample) was directly spread onto a MacConkey and an Ashdown's agar plates to test for the presence of *B.pseudomallei* and to quantify bacterial load per sample. Urine sample was centrifuged at 3,000 g for 5 minutes. 10 µL of urine pellet was streaked to an Ashdown's agar plate. Both plates were incubated at 37 °C for 2 (MacConkey agar) and 4 days (Ashdown's agar), respectively. The culture was inspected daily for colonies of *B. pseudomallei*.

**Respiratory secretion and bodily fluid sample**

Respiratory secretion samples (sputum, tracheal suction and pleural fluid) and bodily fluid samples (pus, tissue specimen, synovial fluid or other fluid) were collected from patients suspected of having melioidosis. Using a sterile loop, a neat sample (10 µL) was plated on blood agar, MacConkey agar and Ashdown's agar plates to test for the presence of *B. pseudomallei* as well as to quantify bacterial load per sample. A neat sample was also inoculated into selective broth. Both the plates and the broth were incubated at 37 °C for 2 days and inspected daily for positive culture. The positive broth culture was subsequently subcultured onto Ashdown's agar plate to identify bacterial species.

**Study 2 Protocol: a diagnostic evaluation of the sensitivity and specificity of CRISPR-BP34 assay**

To improve the melioidosis diagnosis observed in study 1, an alternative CRISPR-BP34 assay was performed on frozen specimens obtained from patients suspected of having melioidosis in the same hospital setting. The results and time taken to process the samples were recorded to enable the head-to-head comparison of the CRISPR-BP34 and the routine culture performance. The study was granted permission to use clinical samples as well as ethical approval from the Ethical Review Board of Sunpasitthiprasong Hospital (029/65C). The study also received ethical exemption from Chiang Mai University (9190/2565). The laboratory protocol was described as below.

**Production of CRISPR-Cas12a**

**Expression and purification of MBP-LbCas12a**

The methods for expression and purification followed the same procedures as described in the previous study[2].

**CRISPR RNA synthesis**

The CRISPR RNA, crBP34, was reported previously[2] and used in this study. The method for crRNA synthesis followed the same procedures as described in the previous study.

**Generation of genetically-modified *E. coli* strain**

This genetically-modified *E. coli* strain was used for spiking in blood and urine samples to determine molecular sensitivity of the CRISPR-Cas12a detection procedure. This strain was created using $\lambda$ Red recombinase-mediated DNA recombination[3] at the lacZ locus in a parental *E. coli* strain, NEB 10-beta (NEB, USA, #C3019H). The target RPA amplicon of the CRISPR-Cas12a was first amplified by PCR using KOD-Plus-Neo (Toyobo, Japan, #KOD-401) and primers containing adapter sequences (primer 200/201). The PCR amplicon was gel purified by TIANgel Midi Purification kit (TIANGEN, China, #4992443) and cloned into pACYCDuet-1 (Novagen, USA, #71147) at NcoI restriction site (ThermoFisher Scientific, USA, #ER0571). The DNA sequences in the plasmid that encompassed the RPA amplicon and Chloramphenicol resistance gene were then PCR amplified from the plasmid (using primer 221/222) and subsequently gel purified. This PCR amplicon represents the DNA that will be inserted into the lacZ locus using $\lambda$ Red recombinase-mediated DNA recombination. This process requires homology arms on both the upstream and downstream of the insertion point. To accomplish this, ~500-bp DNA sequences of the upstream and downstream regions were separately amplified from the genomic DNA of the *E. coli* NEB 10-beta strain using PCR. The two resulting PCR amplicons were then gel purified. It is important to note that some of these primers that were used in these PCR reactions (primer 217/218 and 219/220) contained overlapping sequences with the DNA was intended to be inserted, allowing the three PCR amplicons to be assembled into a targeting cassette in a subsequence PCR reaction (using primer 217/220)[4].

To insert the targeting cassette into the lacZ locus, a modified protocol derived from previous studies was developed[3,5]. Briefly, pKD46 (a gift from Dr. Aiyada Aroonsri, BIOTEC, Thailand), which expresses $\lambda$ Red recombinase enzymes, was electrotransformed into NEB 10-beta *E. coli* using 1.8 kV, 200 Ω, 25 μF parameters. The cells were grown overnight on LB agar (Miller) (Titan, India, #TM376) in the presence of 100 μg/mL Ampicillin (AppliChem, Germany, #A0839) at 30 °C, and a single colony was selected and grewa starter culture. This starter culture was inoculated into 150-mL LB broth (Miller) (Titan, India, #TM406) in the presence of Ampicillin and 10 mM L-arabinose (TCI, Japan, #A0515) to induce expression of the $\lambda$ Red recombinase enzymes. Cells were allowed to grow at 30 °C until $O.D._{600}$ reached 0.6. They were then harvested and washed three times in an ice-cold solution of 10 %v/v glycerol to ensure complete removal of any impurities, and finally they were resuspended in 1 mL of the same solution. 1.0-1.5 μg of the purified targeting cassette was electroporated into 40 μL of these cells. The cells were recovered in LB (Miller) broth at 37°C for 3 hours with gentle agitation and then plated on LB (Miller) agar containing 5 μg/mL Chloramphenicol (AppliChem, Germany, #A1806) overnight at 37°C. All visible colonies were picked and re-streaked onto LB (Miller) agar containing 7 μg/mL Chloramphenicol to remove any contaminant cells. The genomic DNA of the genetically modified *E. coli* was extracted using the GeneJET Genomic DNA Purification Kit (ThermoFisher Scientific, USA, #K0721). The DNA was then subjected to PCR (using primer 198/199), and the amplicons were gel purified and sent for sequencing to confirm the accuracy of the inserted DNA sequence. Correct clones were stored in 20 %v/v glycerol at -80°C.

**CRISPR-BP34 assay**

**Sample preparation**

For urine sample, 5 mL of urine sample was centrifuged at 3,000 g for 5 minutes. The supernatant was carefully removed, and the sample pellet was resuspended in 1 mL of PBS The resuspended sample was then transferred to a new microcentrifuge tube and centrifuged at 10,000 g for 5 minutes. The supernatant was again carefully removed, leaving approximately 20 µL on top of the sample pellet. This sample was then used for the CRISPR-BP34 assay, as outlined in the protocol provided below and in **Supplementary Figure S4**. For blood culture, respiratory secretion samples and other body fluid, a 200-µL aliquot from samples can be used directly. If the sample was viscous, 1 mL of PBS was used to dilute the sample before transferring. This sample was then proceeded to CRISPR-BP34 assay outlined below and in (**Supplementary Figure S5**)

After sample preparation, 200 µL of the clinical sample was aliquoted into a 1.5 mL test tube. We used 200 µL sample volume because of convenience in sample handling; however, a larger volume is also possible to perform this step. This CRISPR-BP34 assay consists of (i) host cell depletion, (ii) DNA extraction, (iii) RPA and (iv) CRISPR reaction and detection

**Host cell depletion**

This step is required for samples that are contaminated with significant amounts of human host cells, for example, blood, hemoculture, sputum and pus. Contamination of host cells' genomic DNA negatively impacts sensitivity and specificity of nucleic acid amplification[6–10]. Since bacterial pathogens contain a rigid cell wall, depletion of host cells' DNA can be simply achieved by selectively lysing the mammalian cells with a mild detergent-containing buffer followed by centrifugation to collect intact bacterial cells before proceeding to DNA extraction. To deplete host cells, an equal volume of 200 µL of a Mammalian Cell Lysis Buffer-1[11] (MCLB-1, containing 2 M $Na_2CO_3$ pH 9.8 and 1% v/v Triton-X 100) was added to the test tube. The test tube was then vigorously vortexed every 15-20 seconds during a 4-minute incubation at room temperature. The mixture was neutralized with a Neutralizing buffer (Tris-HCl pH 4.5), briefly vortexed and centrifuged at 10,000 g for 5 minutes. The supernatant containing DNA and cytoplasmic contents of human cells was carefully discarded without disturbing a bacteria-containing pellet at the bottom. This pellet was then washed with 1 mL PBS, briefly vortexed and centrifuged at 10,000 g for 5 min. This washing step could be repeated one more time if the wash supernatant was still contaminated with host cells' constituents such as blood pigments. The wash supernatant was carefully removed and left behind ~20 µL atop the bacteria-containing pellet. This host cell-depleted sample was then proceeded to the DNA extraction step. Technical details of the host cell depletion step were presented in **Supplementary Figure S3** and **S5**.

**DNA extraction from clinical samples**

While a simple and streamlined methodology for extracting pathogen DNA is preferred, some clinical samples remain complex in nature even after the host cell depletion step. These samples prove difficult to be processed by the simple extraction method. In our protocol, bacterial DNA can be easily extracted by alkaline lysis if the bacteria-containing pellet is less than 10 µL in volume. This method has been successfully applied for urine, whole blood, and hemoculture samples. However, samples such as sputum, pus, and respiratory secretions remain viscous and large in volume even after host cell depletion, necessitating a more thorough extraction method using a commercial spin column. To facilitate sample processing, we have developed pipelines that guide the processing of different clinical samples using easy-to-follow flowcharts, which can be found in **Supplementary Figure S3, S4** and **S5**.

**Alkaline lysis**

This DNA extraction method is a modified version of a method derived from a patent[12], and was used to extract DNA from a bacteria-containing pellet that was less than 10 μL in size. Typically, these pellets were obtained from centrifuged urine samples or host-cell depleted whole blood or hemoculture samples. To perform this step, 2 μL of 550 mM NaOH (i.e. alkaline) was added and mixed with the pellet. This resulted in a final concentration of 50 mM NaOH. The sample was heated at 70 °C for 10 minutes to lyse bacterial cells, after which 2.5 μL of 1 M Tris pH 7.5 was added to neutralize the sample (final Tris concentration ~100 mM). The resulting sample could be used directly in a RPA reaction or stored at -20 °C until needed. Technical details of this step were presented in **Supplementary Figure S3** and **S4**.

**GeneJet spin column**

This extraction method was utilized for extracting DNA from bacterial-containing pellets larger than 10 μL. Such pellets could be derived from centrifuged urine samples, host-cell depleted whole blood, or hemoculture samples. Bodily fluids such as pus, sputum, and respiratory secretions always yield a bacterial-containing pellet larger than 10 μL after the host cell depletion step; therefore, they are always processed using this extraction method. To purify the genomic DNA, the GeneJet Genomic DNA Purification Kit (ThermoFisher Scientific, USA, #K0721) was used with a modified protocol as follows. The pellet was resuspended in 200 μL of PBS, and 200 μL of lysis buffer along with 20 μL of proteinase K were added. The mixture was incubated at 56 °C for 30 minutes. Then, 20 μL of RNase A was added, and the mixture was further incubated at 37 °C for 10 minutes. Next, 400 μL of 50 %v/v ethanol was mixed in, and the resulting mixture was transferred to the spin column and washed following the manufacturer's protocol. The DNA was eluted in only 30 μL of an elution buffer at 56 °C for 2 minutes, before being spun down to a new test tube at 12,000 g for 1 minute. The DNA was stored at -20 °C until use. Technical details of this step were presented in **Supplementary Figure S5**.

**Recombinase polymerase amplification (RPA)**

TwistAmp Basic (TwistDx, USA, #TABAS03KIT) was used for RPA in accordance with the manufacturer's protocols, with modifications as follows. (i) The total volume of an RPA reaction was adjusted to 30 μL, (ii) incubation was performed at 39℃ for 30 minutes, (iii) DNA input was only 2 μL (adding more DNA to the reaction did not necessarily increase sensitivity, as it could contain some inhibitors) and (iv) MgOAc was added last to initiate the reaction. All RPA reactions were stored at -20℃ until used. Technical details of this step were presented in **Supplementary Figure S6**. RPA primers, 148 and 149, were previously reported[2], synthesized and purified by standard desalting by Macrogen (Korea).

**CRISPR reaction and dipstick detection**

CRISPR reactions were performed in 50-μL volume that contained 100 nM crRNA, 200 nM MBP-LbCas12a, 100 nM FAM-biotin probe, 5 μL RPA reaction and 1x HOLMES buffer 1[13] (2 mM spermidine, 40 mM Tris pH 8.5, 6 mM $MgCl_2$, 1 mM DTT, 40 mM glycine, 0.001 %v/v Triton X-100, 0.4 %w/v PEG-20,000). The reaction was incubated at 37 °C for 60 minutes. Thereafter, a lateral flow dipstick (Milenia Biotek, Germany, #MGHD1) was directly dipped into the reaction and allowed to develop for 5 minutes before reading. Technical details of this step were presented in **Supplementary Figure S7**.

**Determining the bacterial loads from clinical specimens**

The number of colony forming units (cfu/mL) of *B. pseudomallei* was estimated from the positive agar plates from hemoculture; genitourinary fluid; respiratory secretion; and pus, tissue and other body fluid samples as part of the laboratory practice. The number of bacterial cells in direct blood samples was aggregated from the former dataset14. Blood collected directly from patients who had not been treated and suspected to have melioidosis was transferred into isolator tubes. The blood was then lysed to release intracellular bacterium, and centrifuged to condense the number of bacterial cells before being plated on blood agar plates. The CFU was counted.

**qPCR validation**

Three sets of qPCR primers, namely 192/193, 270/271, and 274/275, were utilized to detect *B. pseudomallei* genomic DNA in samples that produced inconsistent results between CRISPR-BP34 assay and either standard culture-confirmed assay or hemoculture. These samples were usually positive for the CRISPR assay but negative for the culture methods, likely due to their low sensitivity. The three primer pairs target two different genes in order to increase the likelihood of detection, as a meta analysis indicated that targeting a single gene TTS1 resulted in a qPCR sensitivity of only 77.1%[15]. These three primer pairs are listed in **Supplementary Table S1** and were synthesized and desalted by Macrogen (South Korea). The qPCR reactions were prepared in a 20-µL volume using Maxima SYBR Green/ROX qPCR Master Mix (ThermoFisher Scientific, USA, #K0221), following the manufacturer's instructions. Duplicated qPCR reactions were carried out on the Applied Biosystems 7500 Fast Real-Time PCR System using the following conditions: an initial denaturation step of 95 °C for 10 minutes, followed by 40 cycles of denaturation at 95 °C for 15 seconds, annealing at 61 °C for 30 seconds (for primer pairs 192/193) or 64°C (for primer pairs 270/271) or 62 °C (for primer pairs 274/275), and extension at 72 °C for 30 seconds. Positive detection was determined if any duplicate reaction in any primer pairs yielded a positive result. Standard melting curve analysis was performed at the end of the PCR and compared to that of positive control reactions. ΔRn threshold was automatically calculated by the built-in program.

**Suggestions to mitigate DNA cross-contamination**

Like other nucleic acid amplification assays, such as PCR and LAMP, RPA is also susceptible to the problem of false positives caused by amplicon cross-contamination. To minimize this risk during the RPA step, we suggest implementing the following practices.

i. Assign a dedicated workstation for the RPA step, if possible, and clean the workstation before and after the RPA step
ii. Avoid air turbulence, such as in a biosafety cabinet, when performing this step or when opening an RPA test tube
iii. Use filter pipette tips and have a dedicated disposal bin for pipette tips that have come into contact with the RPA reaction
iv. Even though we did not perform this, RPA and CRISPR reactions can be coupled into a single 'one-pot' reaction[16]. This eliminates the need to open the RPA test tube, but a fluorescent probe and compatible readout must be utilized
v. It is also possible to routinely incorporate deoxyuridine triphosphate (dUTP) during the RPA step and treat DNA samples with Uracil DNA Glycosylase (UDG) before the RPA reaction[17]. UDG cleaves uracil-containing amplicons, thus reducing cross-contamination.

**SUPPLEMENTARY FIGURES**

**Supplementary Figure S1. A setup of a workstation for CRISPR-BP34 assay**

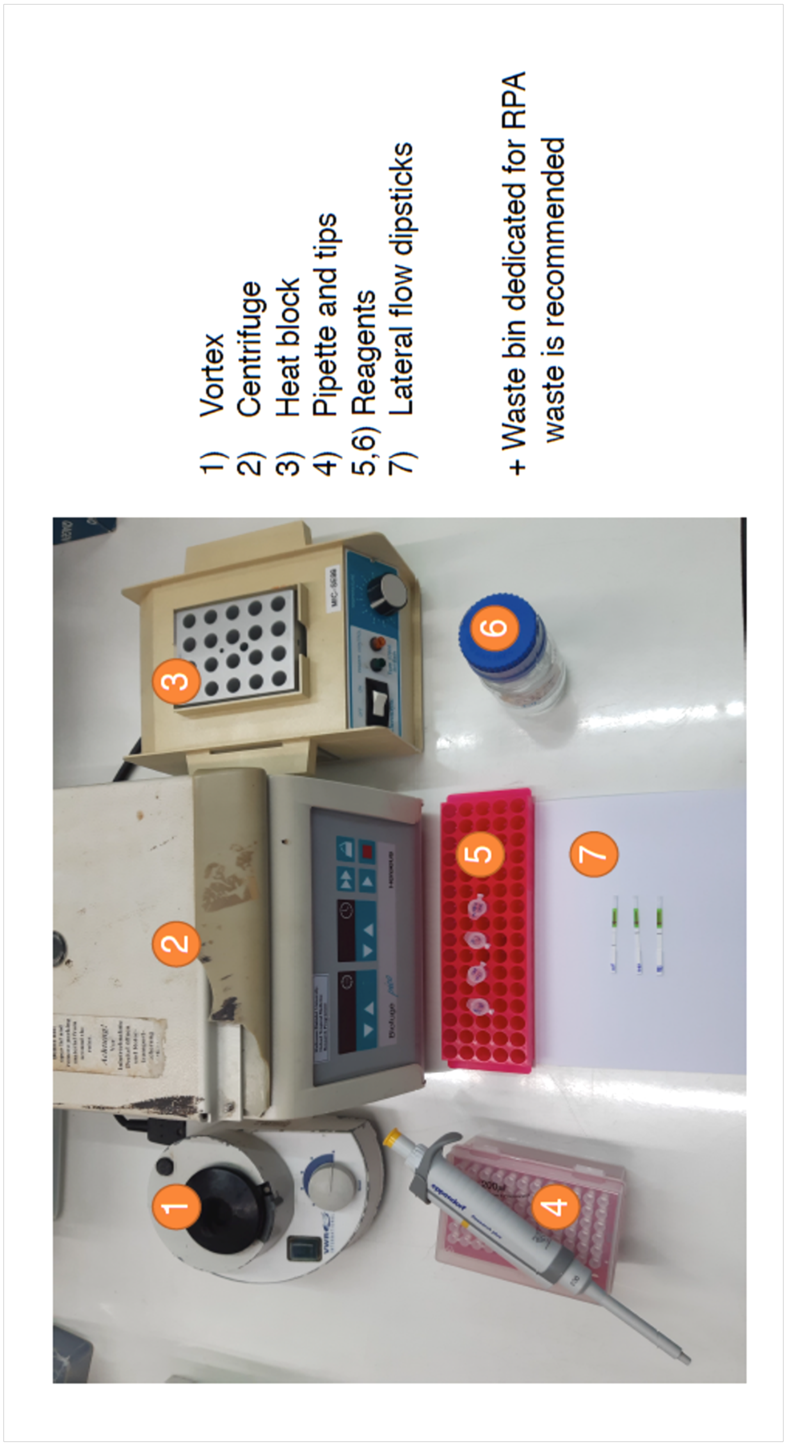

**Supplementary Figure S2. A workflow of CRISPR-BP34 assay**

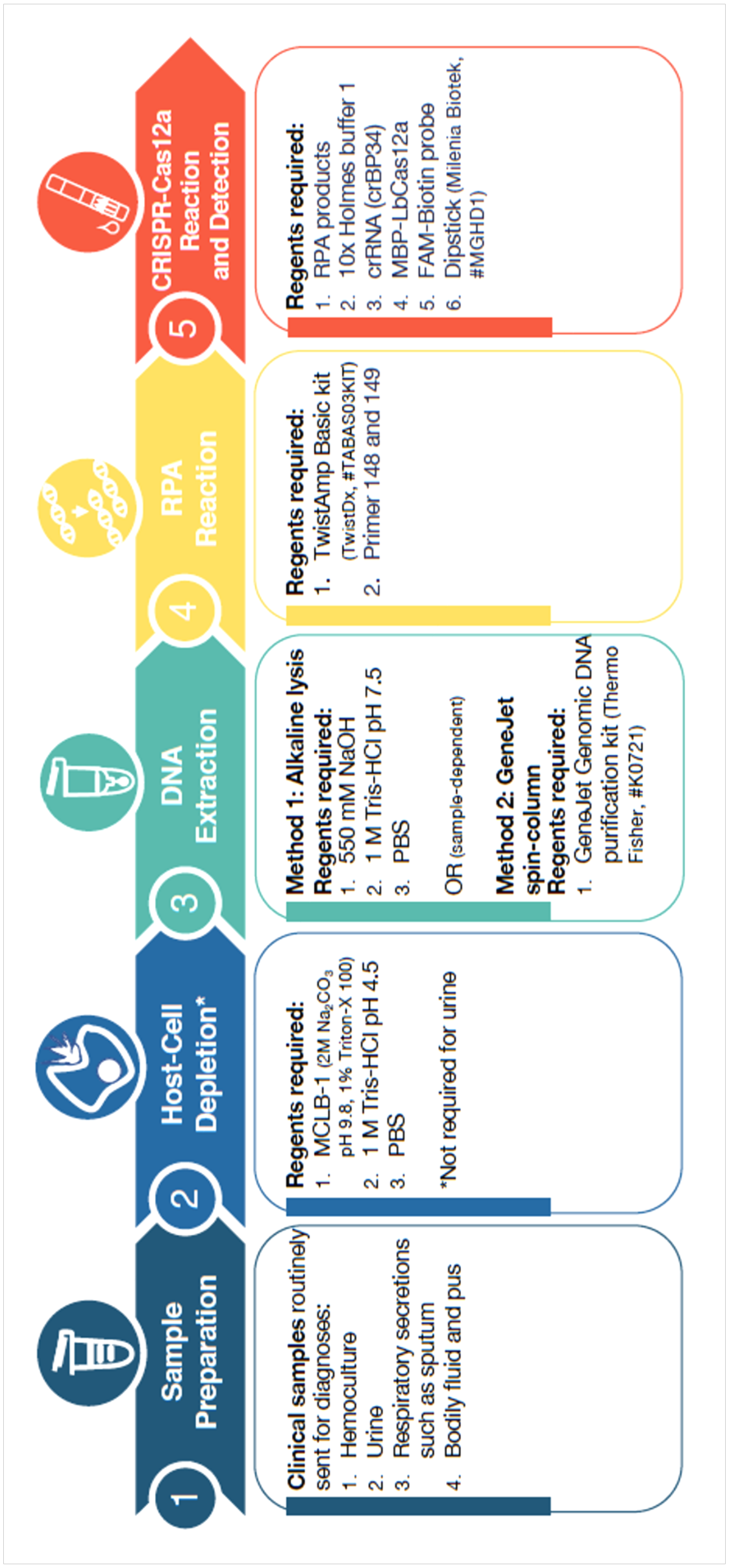

**Supplementary Figure S3. CRISPR-BP34 assay flowchart for hemoculture samples or blood**

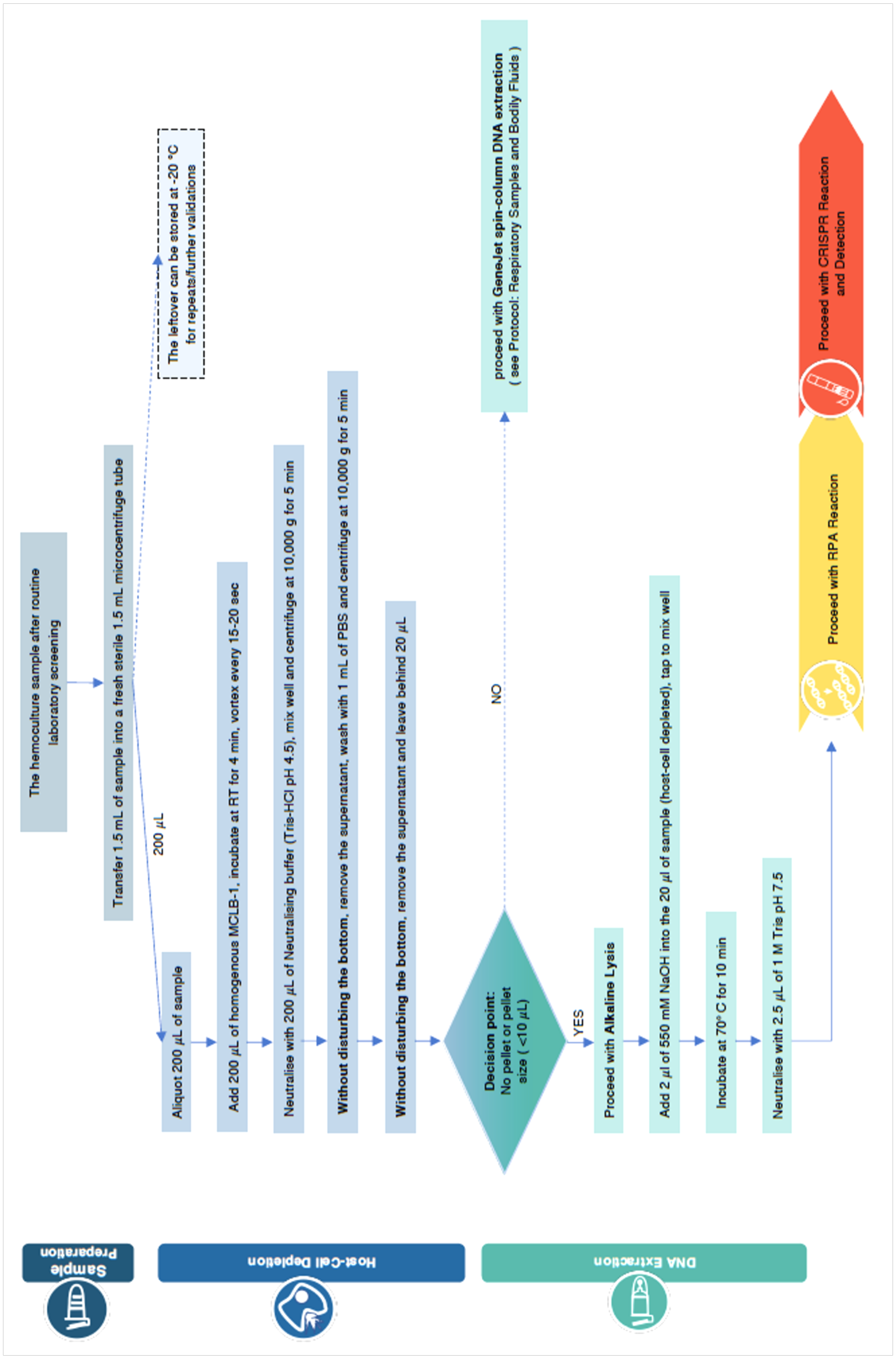

**Supplementary Figure S4. CRISPR-BP34 assay flowchart for genitourinary samples**

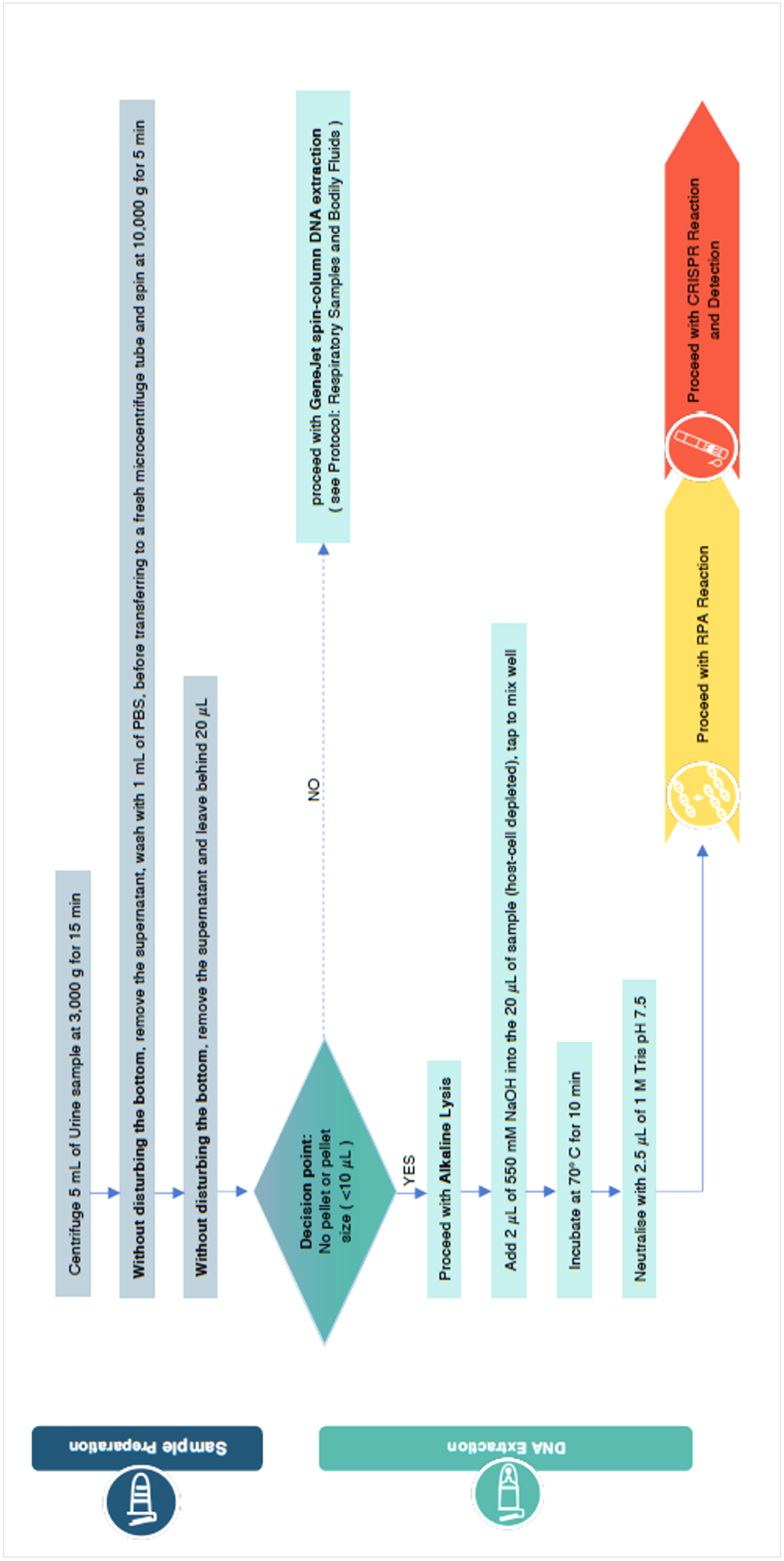

**Supplementary Figure S5. CRISPR-BP34 assay flowchart for bodily fluids** (such as sputum or pus)

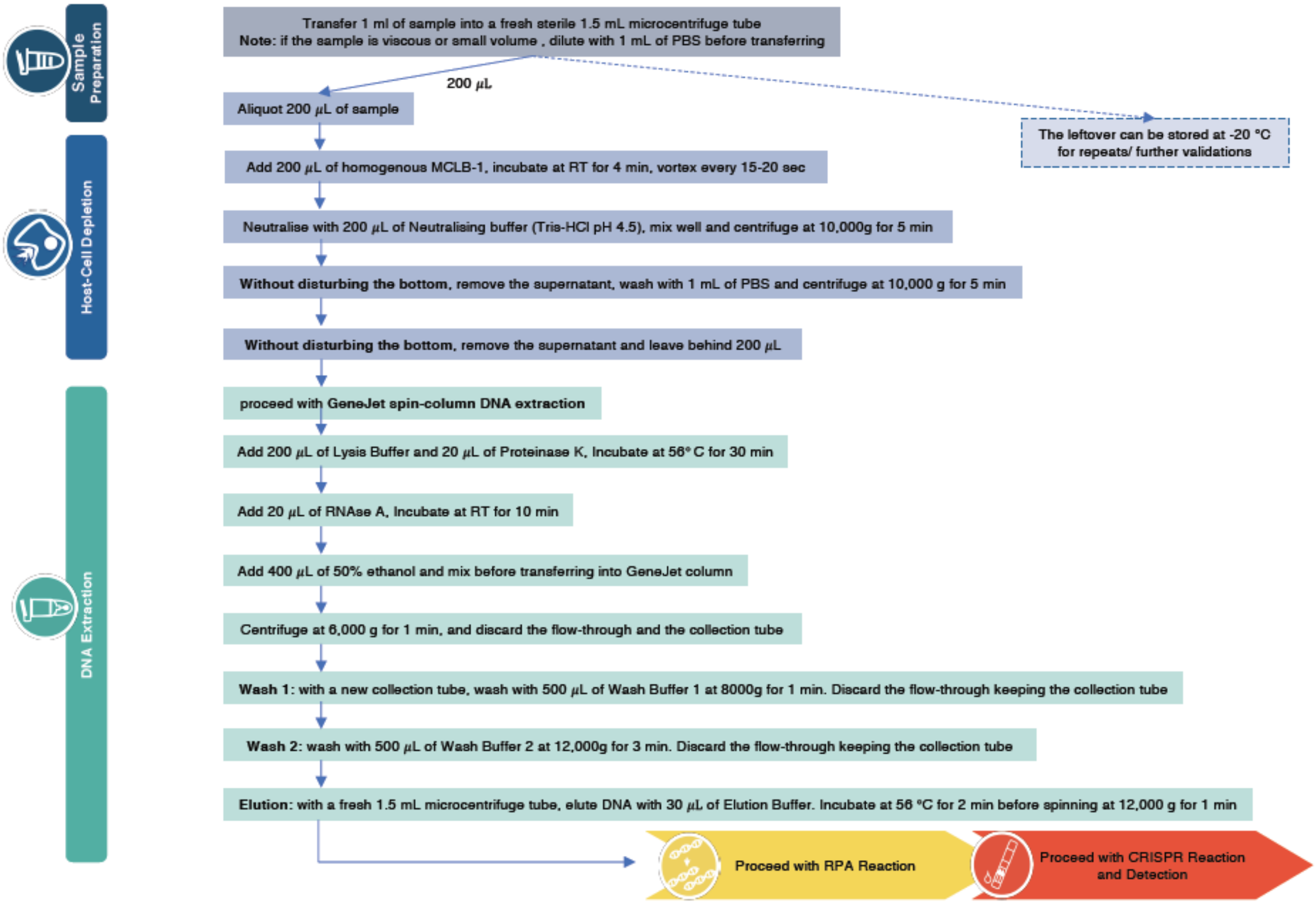

**Supplementary Figure S6. RPA reaction assay flowchart**

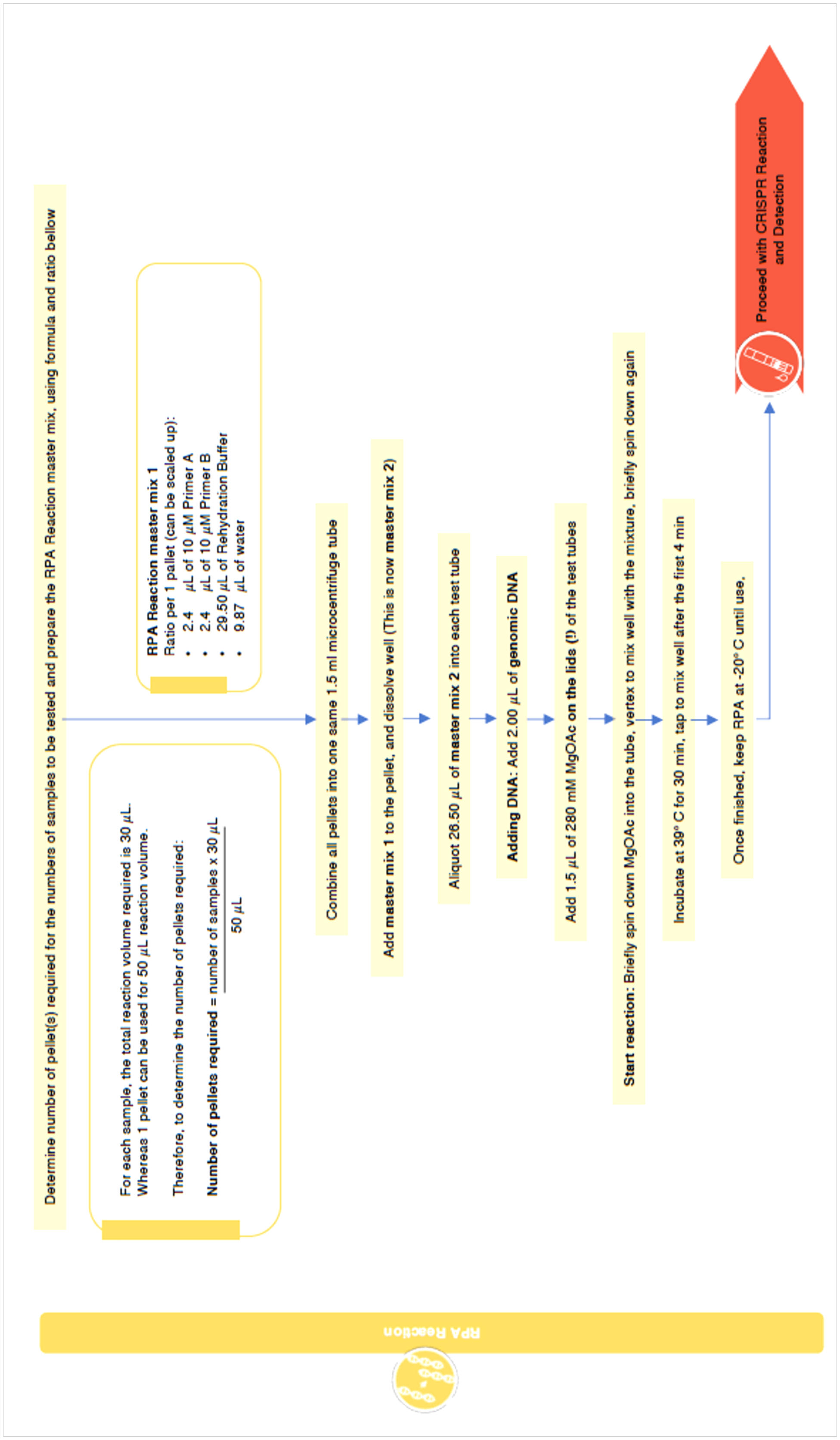

**Supplementary Figure S7. CRISPR reaction and dipstick detection assay flowchart**

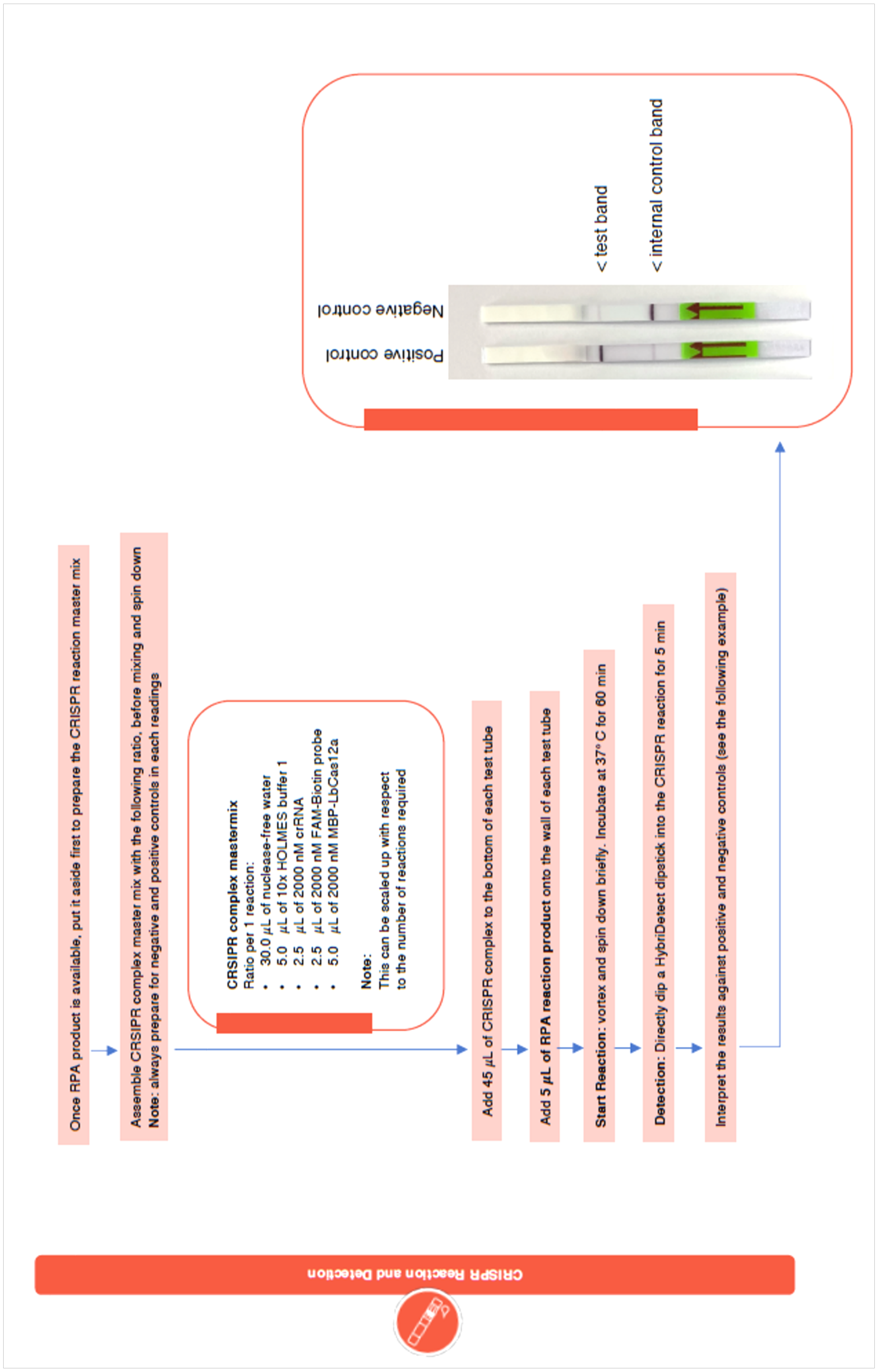

**Supplementary Figure S8. Comparison of qPCR cycle threshold (ct scores) between culture positive and negative samples.**

Scatter boxplots summarise the number of ct scores obtained from qPCR performed to validate culture negative but CRISPR-BP34 positive samples. Each individual dot represents a single test while each boxplot highlights the first quartile, median, and third quartile, respectively. Culture positive (hemoculture) samples were used as a positive control (left). Blank water was used as negative control (results not shown) Hemoculture (pink), urine (orange), respiratory secretion (purple) and pus (blue) with culture negative but CRISPR-BP34 positive results all show significantly higher ct scores. This suggests the presence of *B. pseudomallei* at low bacterial loads which are possibly under the culture limit of detection.

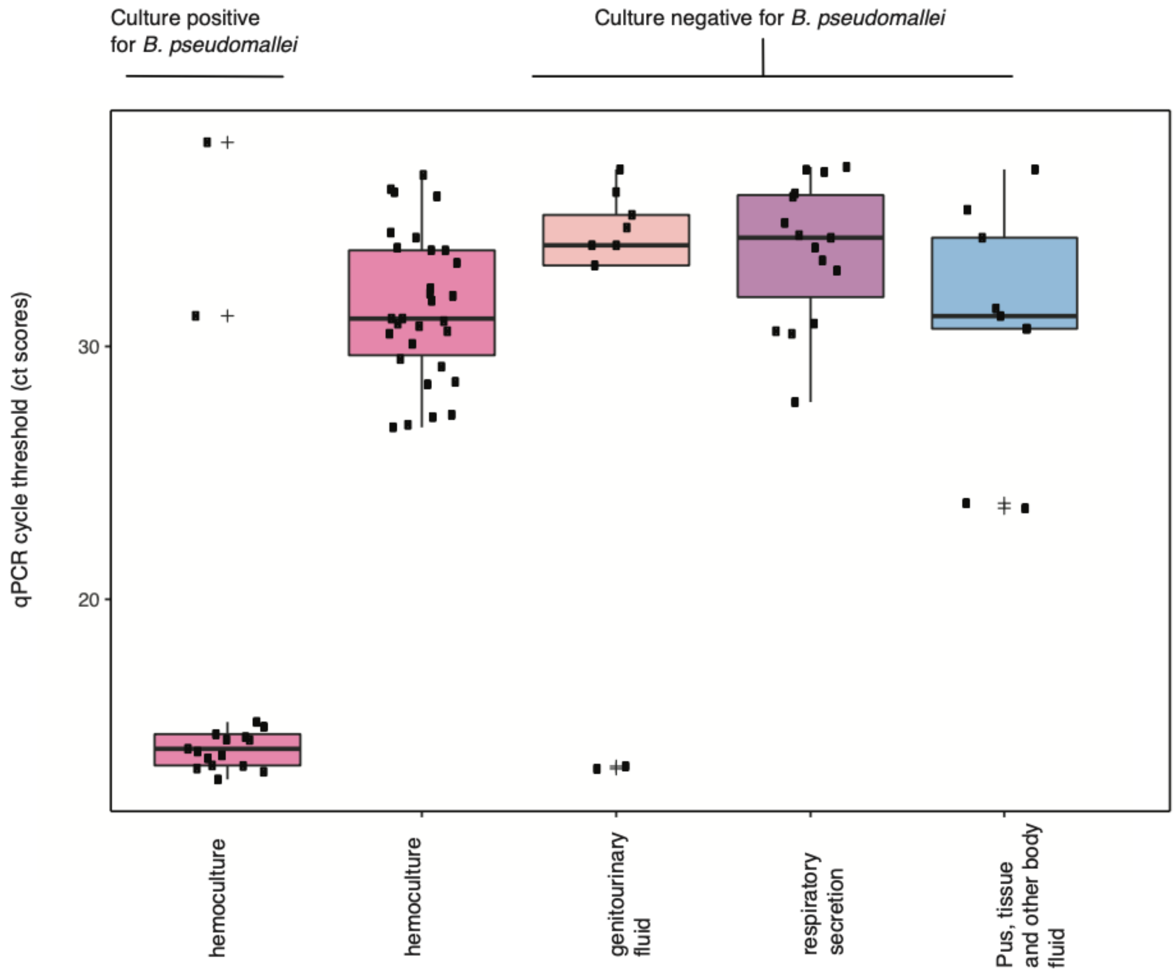

**Supplementary Figure S9. Symptoms reported in melioidosis patients**

An upset plot summarises the clinical manifestations and their intersecting sets of melioidosis patients admitted or referred to Sunpasitthiprasong hospital between October 2019 and December 2022. and their intersecting sets. Major eported symptoms ranged from fever, cough, dyspnea, GI disturbance, urinary symptoms, joint pain, muscle pain, local mass, swollen or abscesses, and weightloss

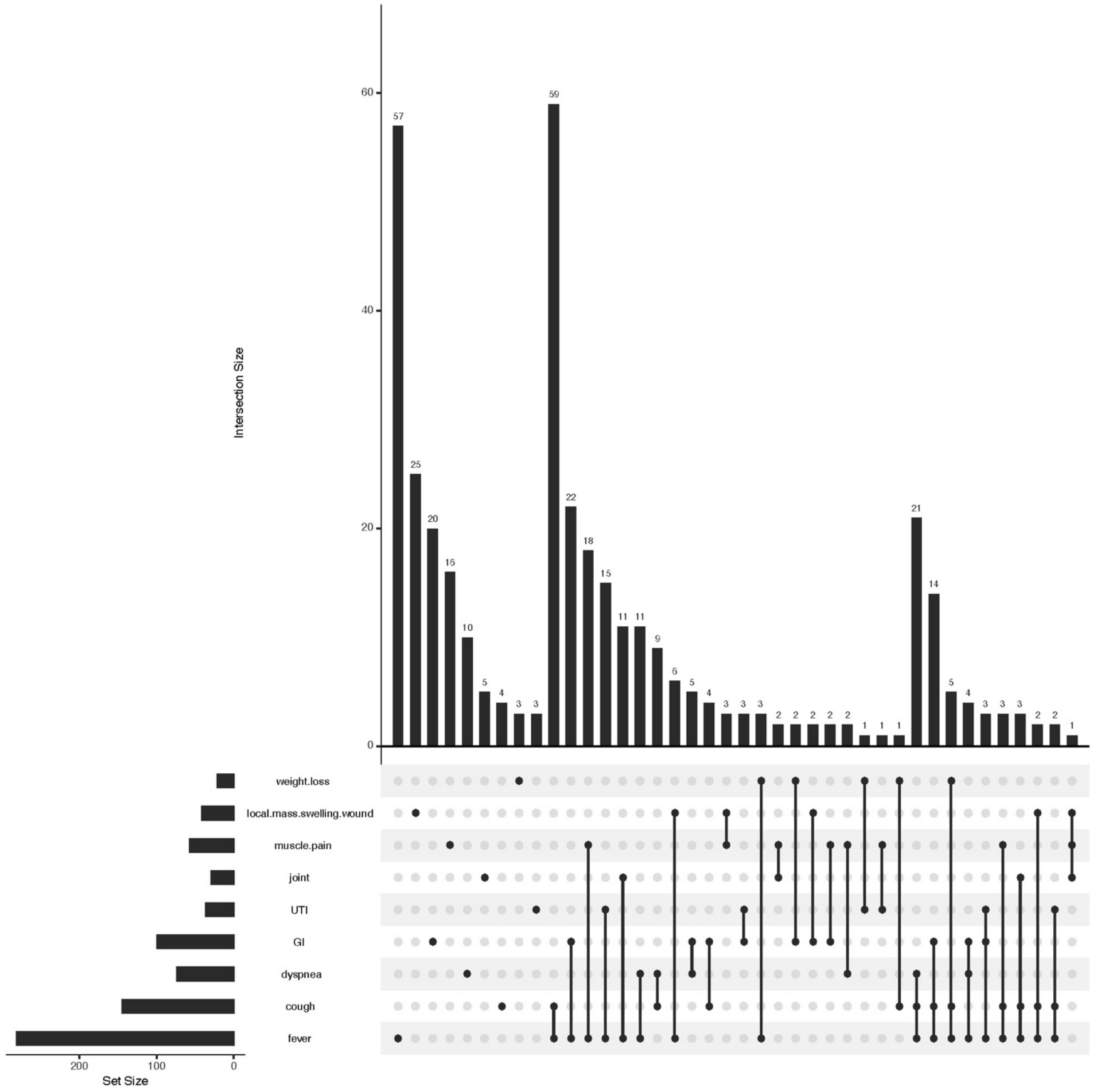

### SUPPLEMENTARY TABLES

#### Supplementary Table S1. A list of primers and oligos used in the study

| Name | Sequences | Usage | Reference |
|---|---|---|---|
| 148 | CAGCATATCATTGTCCGGCGCGAACCATCAAGCTA | RPA primer for crBP34 | [2] |
| 149 | AACTTTTCATTTTCCTGTCAATTCGACTGACCATC | RPA primer for crBP34 | [2] |
| 167 | TAATACGACTCACTATAGGGTAATTTCTACTAAGTGTAGATACTACATACCCACTATTCAGAAAGGAA | Annealing to DNA oligo 168 to create a T7 transcription template for crBP34 crRNA | [2] |
| 168 | TTCCTTTCTGAATAGTGGGTATGTAGTATCTACACTTAGTAGAAATTACCCTATAGTGAGTCGTATTA | Annealing to DNA oligo 167 to create a T7 transcription template for crBP34 crRNA | [2] |
| 192 | CGTCTCTATACTGTCGAGCAATCG | qPCR primer targeting orf2 within putative type III secretion system cluster 1 (TTS1) of *B. pseudomallei* | [18–20] |
| 193 | CGTGCACACCGGTCAGTATC | qPCR primer targeting orf2 within putative type III secretion system cluster 1 (TTS1) of *B. pseudomallei* | [18–20] |
| 198 | ATGCGACTCCTGCATTAGGAA | Amplifying the DNA insert from the genome of *E. coli* NEB 10-beta | This study |
| 199 | TTCGATTATGCGGCCGTGTA | Amplifying the DNA insert from the genome of *E. coli* NEB 10-beta | This study |
| 200 | AAGGATCCATGGCAGCATATCATTGTCCGGCG | Cloning of the RPA amplicon into pACYCDuet-1 | This study |
| 201 | AAGGATCCATGGAACTTTTCATTTTCCTGTCAATT | Cloning of the RPA amplicon into pACYCDuet-1 plasmid | This study |
| 217 | TATGGAAACCGTCGATATTCAGC | Amplifying ~500-bp DNA from the lacZ locus and utilizing it as the upstream homology arm | This study |
| 218 | TTCCTAATGCAGGAGTCGCATAAGTGCACGGCAGATACACTTG | Amplifying ~500-bp DNA from the lacZ locus and utilizing it as the upstream homology arm | This study |
| 219 | GAAGTGATCTTCCGTCACAGGGATCATCGGTCAGACGATTCATT | Amplifying ~500-bp DNA from the lacZ locus and utilizing it as the downstream homology arm | This study |
| 220 | CGAGTTGCGTGACTACCTAC | DNA from the lacZ locus and utilizing it as the downstream homology arm | This study |
| 221 | TTATGCGACTCCTGCATTAGGAA | Amplifying RPA amplicon and Chloramphenicol resistance gene from pACYCDuet-1 plasmid | This study |
| 222 | CCTGTGACGGAAGATCACTTC | Amplifying RPA amplicon and Chloramphenicol resistance gene from pACYCDuet-1 plasmid | This study |

| Name | Sequences | Usage | Reference |
|---|---|---|---|
| 270 | AACACTGACAAGTGGCCCTATGGA | qPCR primer targeting orf11 (BPSS1386) of *B. pseudomallei* | [21] |
| 271 | TCCGATCGGTTTCGAATAACGGGT | qPCR primer targeting orf11 (BPSS1386) of *B. pseudomallei* | [21] |
| 274 | CTTCGGACCGCCTCGAACT | qPCR primer targeting crBP36 locus (BPSS1386 and intergenic region) of *B. pseudomallei* | This study |
| 275 | GCGAGGGTGTCGAACTGATGT | qPCR primer targeting crBP36 locus (BPSS1386 and intergenic region) of *B. pseudomallei* | This study |
| FAM-biotin probe | /56-FAM/TTATT/3Bio/ | Collateral probe | [2,22] |
